# New Gain-of-Function Mutations Prioritize Mechanisms of HER2 Activation

**DOI:** 10.1101/2025.03.03.25323043

**Authors:** Christopher J. Giacoletto, Liz J. Valente, Lancer Brown, Sara Patterson, Rewatee Gokhale, Susan M. Mockus, Wayne W. Grody, Hong-Wen Deng, Jerome I. Rotter, Martin R. Schiller

## Abstract

*ERBB2* (HER2) is a well-studied oncogene with several driver mutations apart from the well-known amplification defect in some breast cancers. We used the GigaAssay to test the functional effect of HER2 missense mutations on its receptor tyrosine kinase function. The GigaAssay is a modular high-throughput one-pot assay system for simultaneously measuring molecular function of thousands of genetic variants at very high accuracy. The activities of 5,886 mutations were classified, significantly more than mutants previously reported. These variants include 112 new *in vitro*, 10 known, and 9 new *in vivo* gain-of-function (GOF) mutations. Many of the GOFs spatially cluster in sequence and structure, supporting the activation mechanisms of heterodimerization with EGFR and release of kinase inhibition by the juxtamembrane domain. Retrospective analysis of patient outcomes from the Genomic Data Commons predicts increased survival with the newly identified HER2 GOF variants.

**Author statement:** *Christopher J. Giacoletto:* Computational methodology, Software, Validation, Formal analysis, Investigation, Writing - Original Draft, Writing – Review and Editing, Visualization; *Liz Valente:* Validation, Formal analysis, Investigation, Writing - Original Draft, Writing – Review and Editing, Visualization; *Lancer Brown:* Validation, Formal analysis, Investigation, Writing - Original Draft, Writing – Review and Editing, Visualization; *Sara Patterson*; Writing - Review and Editing*; Rewatee Gokhale*; Writing – Review and Editing; *Susan Mockus:* Writing – Review and Editing; *Wayne Grody* Writing – Review and Editing; *Hong Wen Deng:* Writing – Review and Editing; Writing – Review and Editing; *Jerome I. Rotter:* Writing – Review and Editing; *Martin R. Schiller*: Conceptualization, Methodology, Validation, Formal analysis, Resources, Data Curation, Writing - Original Draft, Writing – Review and Editing, Visualization, Supervision, Project Administration, Funding acquisition. The experimental and bioinformatic investigation was performed at Heligenics.

## INTRODUCTION

HER2 is the receptor produced from the *ERBB2* gene and is one of four family members (HER1-HER4) in humans. HER1 (more commonly known as EGFR), HER2, and HER4 each possess an extracellular ligand-binding domain, a transmembrane domain, a juxtamembrane (JM) domain, a catalytic protein tyrosine kinase (TK) domain, and C-terminal tail. While EGFR is activated by multiple ligands, HER3 and HER4 are receptors for Neuregulins (NRG1 and NRG2, respectively). HER2 does not have a known ligand, and HER3 lacks an active TK domain. These receptors can each homodimerize and heterodimerize with other family members, enabling HER2 and HER3 to act as co-receptors, despite lacking either prototypical ligand binding or TK domains. The HER subfamily, like most other Receptor Tyrosine Kinase (RTK)s, signal through the MAPK and PI3K/Akt pathways to stimulate transcriptional responses and drive cell proliferation.[1]

HER family copy number and missense variants stimulate cell proliferation and are common cancer driver alterations. HER2 is a proto-oncogene that is overexpressed and constitutively activated in various human cancers and the prevalence of its deleterious alleles is similar to that of EGFR.[2] Cancers such as bladder carcinoma, esophageal, endometrial carcinoma, melanoma, and cholangiocarcinoma have HER2 variant frequencies >5%.[2,3] In breast cancer, HER2 amplification (present in 25-30% of cases) is a key marker for one of the three major types of somatic breast cancer (HER2^+^, ER^+^, PR^+^). In contrast, triple-negative breast cancer lacks the overexpression of the HER2, estrogen receptors (ER), and progesterone receptors (PR) markers. HER2^+^ breast cancer is associated with oncogenic driver mutations in HER2, but there have also been reports of HER2 mutations in HER2-negative breast cancers.[4–6]

There are 14 well-characterized oncogenic variants located in the JM and TK domains (Q679L, Q709L, L726F, G776S, L755P, L755S, A775_G776insYVMA, D769H, D769Y, V777L, T798M, V842I, T862A, and H878Y).[3,7–12] These variants were identified and characterized by several different research groups using different types of methods, including associations with human outcomes, cause-effect studies in animal models, cell physiology experiments, and biochemical analyses.[8] Since the variants were generally identified and characterized by different groups and with diverse methodologies, there is a lack of uniformity. This lack of reliability of the variant effect may impede the development of diagnostics and precision therapies.

In our previous work on the HIV Tat driven transcription, the GigaAssay technology achieved 94% accuracy. This performance was rigorously assessed by three independent measures: comparison against activity classification from blinded stable cells lines, benchmarking against established data in the literature, and analysis of random nonsense variants, some expected to abolish and some expected to retain activity.[13] This multifaceted evaluation strategy ensured a comprehensive assessment of the ability of the GigaAssay to distinguish mutants with wild type (WT) or loss-of-function (LOF) transcriptional activity.

In this study, we developed a novel GigaAssay to identify *in vitro* GOF mutations in the well-known oncogene, HER2. All mutants were assayed under identical conditions, and we achieved an accuracy comparable to that of the previous GigaAssay analysis of Tat. The GigaAssay measured phosphorylation levels for a key site in the C-terminus (Y1248), which is phosphorylated upon HER2 activation (pHER2). This site is a commonly accepted marker of HER2 kinase activity used in many studies.[14,15] Activity was assessed with a high-throughput GigaAssay measuring pHER2 immunostaining and by flow cytometry of stable cell lines expressing doxycycline (DOX)-inducible versions of these variants **(Fig. 1A, B)**.

**Figure 1:**
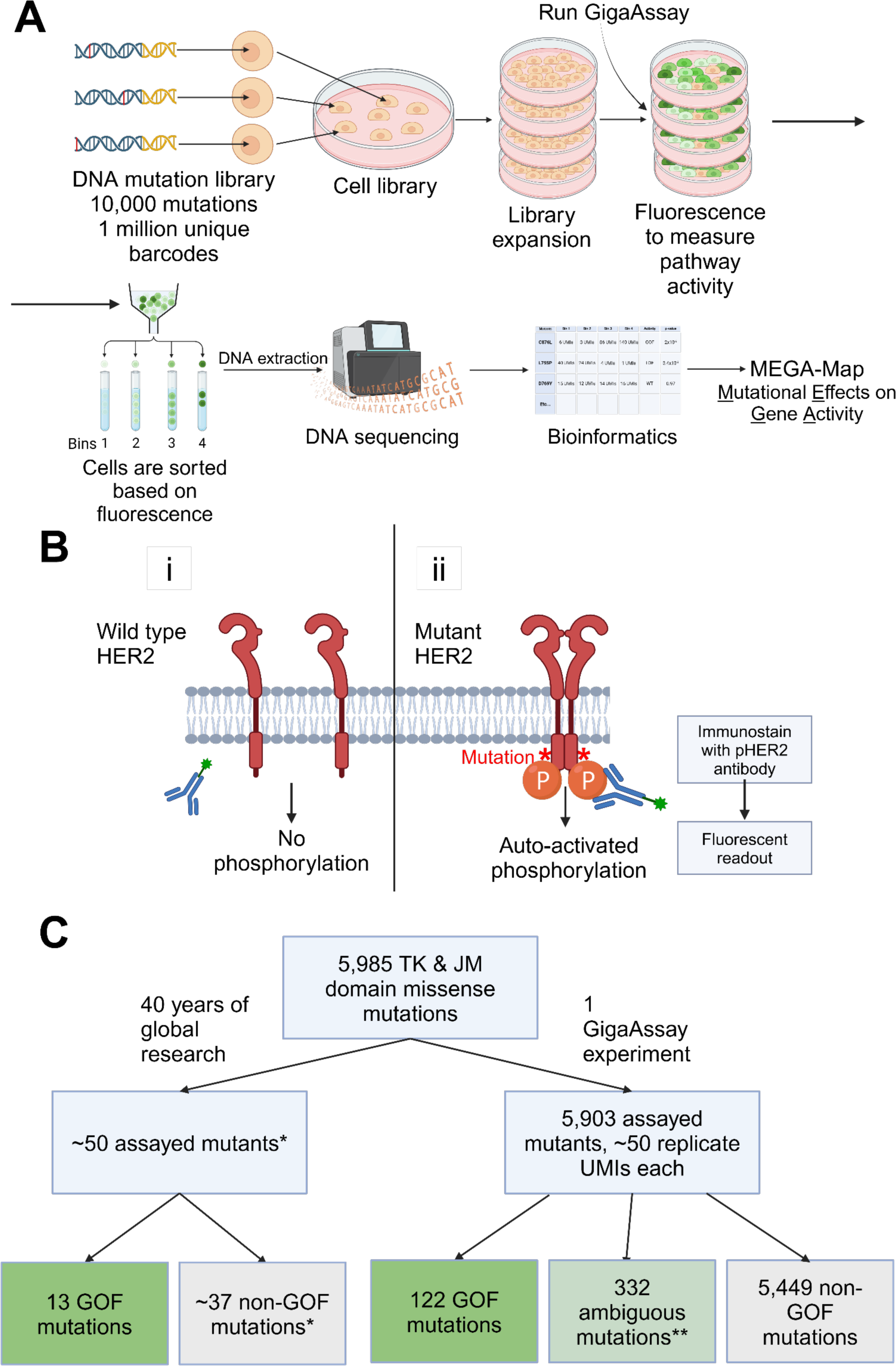
pHER2 GigaAssay architecture. **A.** A flow diagram of the GigaAssay. Designed DNA mutations are transduced into a cell library. Cells are allowed to proliferate, then cells are assayed for HER2 phosphorylation. Cells are sorted into 4 bins using flow cytometry. DNA is extracted from the cells, and sequencing of the 4 bins is done. Bioinformatics and downstream analytics determine the functional activity of all mutants by quantifying the distribution of a mutant’s representation across the 4 bins. **B.** A cartoon representation of the fluorescent readout of pHER2 activity via immunostaining of pY1248 antibody as a surrogate for auto-activated phosphorylation of HER2. **C.** A flow diagram showing classification of mutants in the HER2 TK and JM domains. The left path denotes literature mutants while the right path has GigaAssay mutants. A single asterisk indicates data approximated from a literature review. Double asterisks indicate mutants which have one GOF call and one non-GOF call in technical replicates. Boxes colored green are for GOF mutations and light green is for mutants in which only one of two technical replicates were classified as GOF.

We measured the activity of 5,886 mutations - 100-fold more than previously reported (**Fig. 1C**). We extended the 14 selected GOFs with 112 new *in vitro* GOFs, including 9 new *in vivo* GOF mutations. The HER2 *in vitro* GOFs are clustered by position, physiochemical properties of amino acid sidechains, and by certain molecular functions. We then identified the subset of these mutations that overlap with those reported in various tumor types.

## 2. Results

### 2.1 GigaAssay design

We previously developed a new high-throughput assay system called the GigaAssay that measures the functional activity and effect of thousands of gene mutants in a single experiment. The GigaAssay was used for two experiments examining the Tat-driven transcriptional activities of all possible single amino acid substitutions in two different cell lines.[13,16] For each mutant, the activities were measured in ∼100 unique molecular identify (UMI)-barcoded single cells. This approach achieved a ∼94% accuracy measured by three independent approaches, all of which yielded consistent results.[13,16] Detailed descriptions of the experimental methods and bioinformatics pipelines used in these studies can be found in the previous reports.[13,16]

The HER2 tyrosine kinase receptor can be activated through both ligand-dependent and ligand-independent mechanisms. HER2 homodimers are not thought to bind ligand. Typically, HER2 heterodimerizes with other ErbB family members like Her1 (also called Epidermal Growth Factor Receptor [EGFR]), HER3, or HER4.[17] HER2 tyrosine kinase activity is activated by the epidermal growth factor (EGF)-like ligands such as EGF, TGFα, amphiregulin, and others for EGFR heterodimers and neuregulins for HER3 and HER4 heterodimers with HER2. However, HER2 can also drive signaling and cell proliferation in a ligand-independent manner, particularly in transformed cells harboring HER2 gain-of-function (GOF) mutations. To further explore this ligand-independent activation, a new GigaAssay was developed. This assay was specifically designed to measure ligand-free HER2 phosphorylation at Y1248, a site commonly used as a surrogate marker of HER2-driven signaling activation.

Doxycycline-inducible HEK-293T cells, which express all four Her receptors, provide a suitable model for testing the effects of exogenous HER2 mutants. A GigaAssay was established to measure ligand-free pHER2 phosphorylation at Y1248, a marker of HER2 activation. The assay was optimized using well-characterized HER2 variants exhibiting WT and GOF activity to maximize the dynamic range of the assay. By leveraging the Tet response element (TRE) to control HER2 variant expression, this system reduces artifacts associated with unbalanced growth and variant representation within the cell library. These artifacts can arise from the clonal out-growth of fast-growing clones within the pooled variant library during its preparation. Inducible expression helps to ensure a more balanced representation of variants and reduces the potential for growth-related biases in the assay results.

A saturation mutagenesis gene mutation library (GML) containing HER2 mutants was constructed. The library was subcloned into a lentiviral expression plasmid with each cDNA molecule uniquely tagged with a unique molecular identifier (UMI)-barcode in the 3’ untranslated region (UTR). The resulting lentiviral library was subsequently tested with a GigaAssay by transducing cells with a low multiplicity of infection (MOI) of 0.1 to ensure a single variant integrates in each cell. Following infection, cells were subjected to selection using a poison encoded by the resistance marker. This selection process eliminates cells that did not successfully integrate the lentiviral DNA, such that that the vast majority of remaining cells express a specific HER2 mutant.

After selection, HER2 variant expression was induced by treating cells with 100 ng/mL doxycycline for 3 days. Daily replacement of media with DOX ensured consistent expression levels throughout the experiment. Cells were then harvested, fixed and immunostained with two antibodies: an anti-HER2 monoclonal antibody to detect overall HER2 expression and an antibody targeting pHER2(Y1248) (a marker of HER2 activation) to assess the level of HER2 phosphorylation at Y1248.[18] Cells expressing high levels of HER2 were then sorted into pools using fluorescence-activated cell sorting (FACS) based on the intensity of pHER2(Y1248) immunostaining. This sorting strategy allowed for the separation of cell populations with varying levels of HER2 activation, enabling classification of different HER2 variants based on their ability to induce downstream signaling in the absence of ligand.

To identify HER2 GOF mutations, genomic DNA was extracted from each sorted cell pool, which represents populations of cells exhibiting varying levels of HER2 activation based on pHER2(Y1248) immunostaining intensity. Targeted sequencing was then performed on the UMI-barcode region of the integrated lentiviral cDNA. UMI-barcode counts within each pool were used to generate an activation score for each HER2 cDNA molecule. This score reflects the abundance of each variant in pools, in which the pools were sorted by their level of HER2 activation. To identify GOF mutations, populations of cells expressing the same HER2 variant were compared to a population expressing WT HER2 using their respective UMI-barcode counts. A right-tailed t-test was used to determine if the activation scores of the variant populations were statistically greater than the WT population. Variants exhibiting significantly higher activation scores than WT HER2, as indicated by a significant p-value, were classified as *in vitro* GOF mutations. Mutants that did not show a statistically significant difference from WT were categorized as having non-GOF activity. The WT HER2 used in these experiments was a codon-optimized version designed for high expression such that activity can be determined in the presence of low endogenous HER2 expression. Although not explicitly shown in the data, phosphorylation levels of this codon-optimized WT HER2 did not significantly differ from the reference WT HER2 sequence.

### 2.2 Assessment of well-characterized variants with a pHER2 flow cytometry assay

To test the accuracy of the GigaAssay, a benchmark GOF dataset was created using 14 previously characterized GOF mutants and 7 previously known WT mutant classifications from the literature and the Cancer Knowledgebase.[19] These 21 mutants were introduced into *ERBB2* cDNA lentiviral expression vectors and transduced into reporter cells to generate stable cell lines. Clonal lines were immunostained with a phospho-HER2 (pY1248) antibody and HER phosphorylation levels were measured by flow cytometry. The mutants included GOF mutants Q679L, Q709L, L726F, L755P, L755S, A775_G776insYVMA, G776S, D769H, D769Y, V777L, T798M, V842I, T862A, and H878Y, and 7 mutants with non-GOF activity (Q680R, K753A, K753M, S760A, D845A, D845N, and R868W) as reported from the CIVIC or Cancer Knowledgebase databases.[20–22]

The flow cytometry analysis confirmed an accuracy of 85.7%. This includes 11 of 14 *in vitro* mutants which displayed expected *in vitro* GOF activity, and 7 mutants exhibited expected WT or non-GOF activity, aligning with previously reported HER2 activity.

However, three oncogenic mutations had uncertainty regarding their hyperphosphorylation:

- L726F displayed low basal phosphorylation of HER2, similar to cells expressing WT HER2, consistent with published reports.[23]
- H878Y and Q679L exhibited only a small increase in pHER2 compared to cells with WT HER2. These mutants are reported to show cell-specific HER2 hyper-phosphorylation.[24–26]

One surprising aspect of the benchmark GOF mutants is that they differ in their degree of activity and behavior. While L755S, L755P, and T862A display relatively higher phosphorylation levels similar to the A775_G776insYVMA indel mutation, H878Y and Q679L have only a modest increase in phosphorylation compared to WT HER2.[22] The variability in phosphorylation levels observed in these mutants highlights the complexity of HER2 signal and the potential for diverse mechanism of oncogenic activation.

The GigaAssay offers a unique advantage in evaluating relative outcomes due to its ability to simultaneously measure the functional activity for approximately 50 UMI-barcodes for each variant. These activity distributions can be compared to WT HER2, well characterized mutants, variants of uncertain significance (VUSs), and each other. For example, the GOF mutants V777L, T798M, and T862A exhibited a distinct activity distribution pattern with fewer UMI-barcoded cells having lower activities of “3” or below and a greater proportion of UMI-barcodes showing a maximal signaling score of “4”.[22] This observation is consistent with the higher phosphorylation levels observed for T862A in the flow cytometry assay, which is likely attributed to its location in the ATP binding site.[27] In contrast, L726F, H878Y and Q679L had profiles that were similar to WT (grey bars) and other well characterized mutants with WT-like activity. This finding aligns both with existing literature and their flow cytometry hypophosphorylation profiles observed in the flow cytometry assay.[22]

We considered including L726F in the benchmark low-phosphorylation mutant because it is reported in the literature as under-phosphorylated on HER2, but hyperactive for Erk1/2 as a GOF mutant.[23] The L726F mutant was ultimately excluded as a benchmark mutation for evaluating the GigaAssay because its reported hypophosphorylation was not consistent with oncogenicity being driven by the other GOF mutants. H878Y and Q679L were also excluded from the benchmark dataset because of their ambiguous nature, as they displayed variable phosphorylation levels across different cell lines.[26,28] These three variants may be oncogenic through alternative mechanisms and therefore were excluded from accuracy and performance analyses.[22] The remaining 10 hyperphosphorylated mutants and 7 WT-level mutants constituted a benchmark set to test the accuracy and performance of the GigaAssay results with comparative approaches. It should be noted that the A775_G776insYVMA benchmark GOF was not included in the GigaAssay experiment, as it consists of an indel. The GigaAssay was designed for single amino acid polymorphisms.

The GigaAssay demonstrated high accuracy as evidenced by a prior proof-of-concept study of the HIV Tat transcription factor. In that study, accuracy was assessed with three independent methods across two cell lines, resulting in an average accuracy of 94%. The GigaAssay results reported in Giacoletto et al. has a statistical accuracy of 85%.[22] **However, upon exclusion of the aforementioned mutants having reduced phosphorylation verified by flow cytometry, this accuracy increases to 100%.**This allows us to evaluate a GigaAssay saturation mutagenesis experiment to potentially identify new GOF mutations.

Performance of the pHER2 (Y1248) immunostaining GigaAssay was evaluated by comparing its results to a benchmark dataset, which included a set of well-characterized 10 hyperphosphorylated mutants (excluding A775_G776insYVMA) and 7 non-GOF mutants. Notably, the GigaAssay accurately classified all benchmark mutants, demonstrating 100% accuracy and 100% sensitivity (n =17). Cells expressing GOF mutants had flow cytometry profiles similar to those observed with the A775_G776insYVMA mutant, a well-characterized GOF mutant.[22,29] Seven mutants with known WT activity also displayed flow profiles consistent with WT HER2 in the GigaAssay.[30] Furthermore, 130 of 131 nonsense variants were correctly classified, with one mutant having ambiguous classifications among two technical replicates, consistent with a low false positive rate.[30] The GigaAssay results herein indicate its robustness and reliability as a high-throughput assay system for assessing HER2 variants.

### 2.3 Discovery of new HER2 GOF mutants with the GigaAssay

The GigaAssay confirmed the activity of the benchmark mutations with scores clustering by GOF or non-GOF activity (**Fig. 2A**).[22] Since the pHER2 (Y1248) GigaAssay had 100% accuracy, we assessed whether any of the 5,886 variants are GOF mutants. 450 mutants were classified as GOF mutants in at least one technical replicate, based on a one tailed t-test.

**Figure 2:**
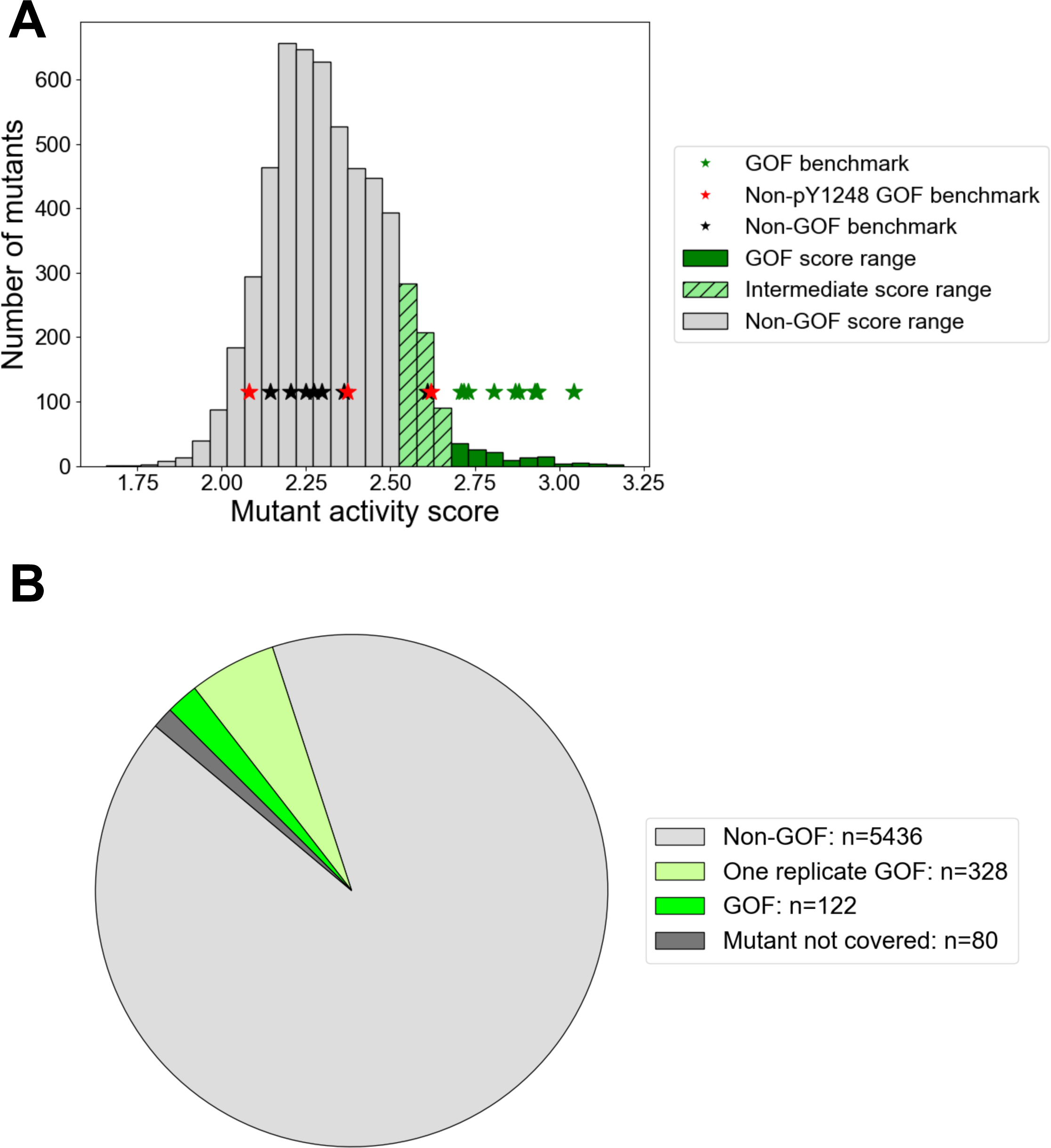
Functional classification and comparison of mutants in replicate GigaAssay. **A.** A histogram of activity scores of all mutants in the GigaAssay. Gray bars are activity scores at, or below that for WT. Green bars indicate scores that are greater than WT. Hatched bars have mutants with scores that could be classified as non-GOF or WT. Green stars indicate scores of the 10 benchmark GOF mutants. Red stars indicate the non-pY1248 GOF benchmark mutations. Black stars indicate activity scores for non-GOF benchmark mutations. **B.** A pie graph showing perecentage s of classified HER2 mutants in technical replicates 1 and 2 of the GigaAssay based on statistical tests. The green slice indicates the percentage of mutants with a GOF classification across both replicates; grey indicates the percentage of mutants with non-GOF classification across both replicates; light green indicates the percentage of mutants with one of two GOF classification call across replicates; an dark gray indicates the small percentage of mutants that lacked sufficient data in at least one replicate to produce a classificaiton.

Of the 5,886 variants, the classification of 94.4% of the mutants replicated in two technical replicates, with only 5.6% having only one measurement or an ambiguous classification. Of the 5,558 mutants that replicated, 122 (2.2%) were *in vitro* GOF in both replicates (2.2%), which includes *all in vitro GOF* benchmark variants (**Fig. 2B**) [22]; the rest were non-GOFs. This is a substantial 10-fold increase in identified GOFs. The 328 mutants with inconsistent classifications, due to lack of statistical congruence, are also potentially GOFs, but will require further testing in orthogonal assays. For the remainder of the paper, we define GOF mutations as the subset identified in both technical replicates, reflecting a more conservative interpretation. In conclusion, this GigaAssay demonstrates 100% accuracy, identifies a 10-fold increase in HER2 GOF mutants, and exhibits high reproducibility.

The mutations, their signaling scores, and classification were plotted on heatmaps (**Fig. 3A, B**) to reveal clustering in the primary sequence. Replicate GigaAssay results are provided in **Supplementary Figs. S1** and **S2.** Heatmaps showing the p-value, number of reads, and UMI-barcodes associated for each mutant are included in **Supplementary Figs. S3-S8**. Several new GOF positions were identified in or around previously known GOF mutations. Notably, this type of validation is rarely observed in other types of high-throughput screens, underscoring the advantages of the GigaAssay.

**Figure 3:**
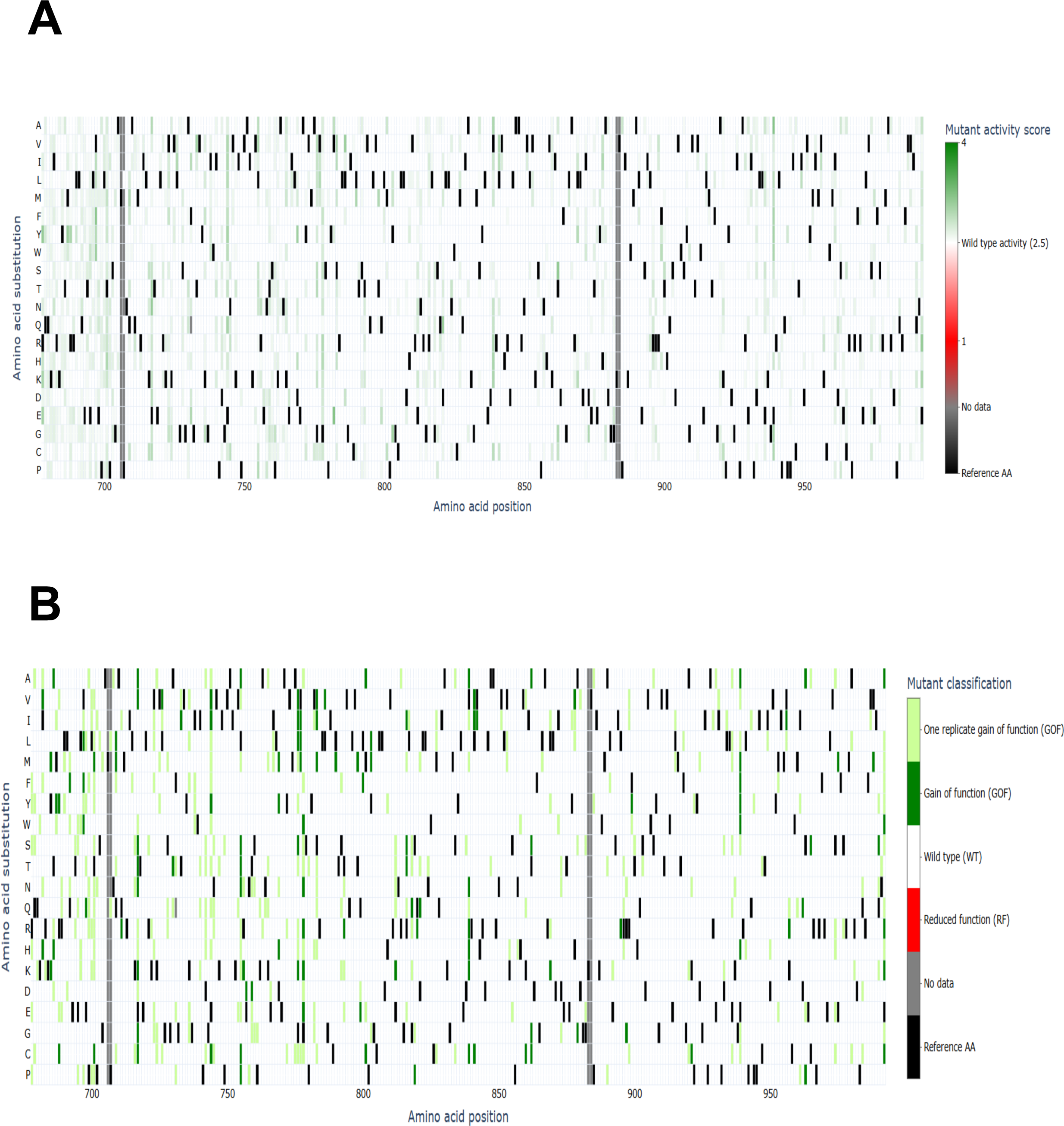
GigaAssay saturation mutagenesis results. Each possible single amino acid substitution is represented by an individual box in the grid. The alternate amino acid is represented on the Y-axis, while the position is represented on the X-axis. The color of the cell indicates the functional activity of the mutation as indicated by the color key. Black squares are the reference sequence, and gray boxes have no data. **A.** The shade of green in the boxes indicate the level of functional activity of the HER2 mutant. **B.** The color of the boxes indicates the classification of the mutants based upon statistical analysis. Data are from replicate 1. Corresponding figures for replicate 2 are in **Supplementary Figs. 1 & 2**.

### 2.4 Structure - Function of TK domain

Excellent studies cover the structure of HER2, as well as mechanisms of action, protein-protein interactions, and quaternary structure interactions.[8,14,15,23,30,31] Multiple intermolecular and intramolecular interactions play key roles in regulating and activating the TK. However, prior to the GigaAssay, the relative significance of these structure/function relationships in oncology were unclear. We describe how newly identified GOF mutations can help prioritize the roles of structure-function elements and regulatory mechanisms (**Fig. 4**).

**Figure 4:**
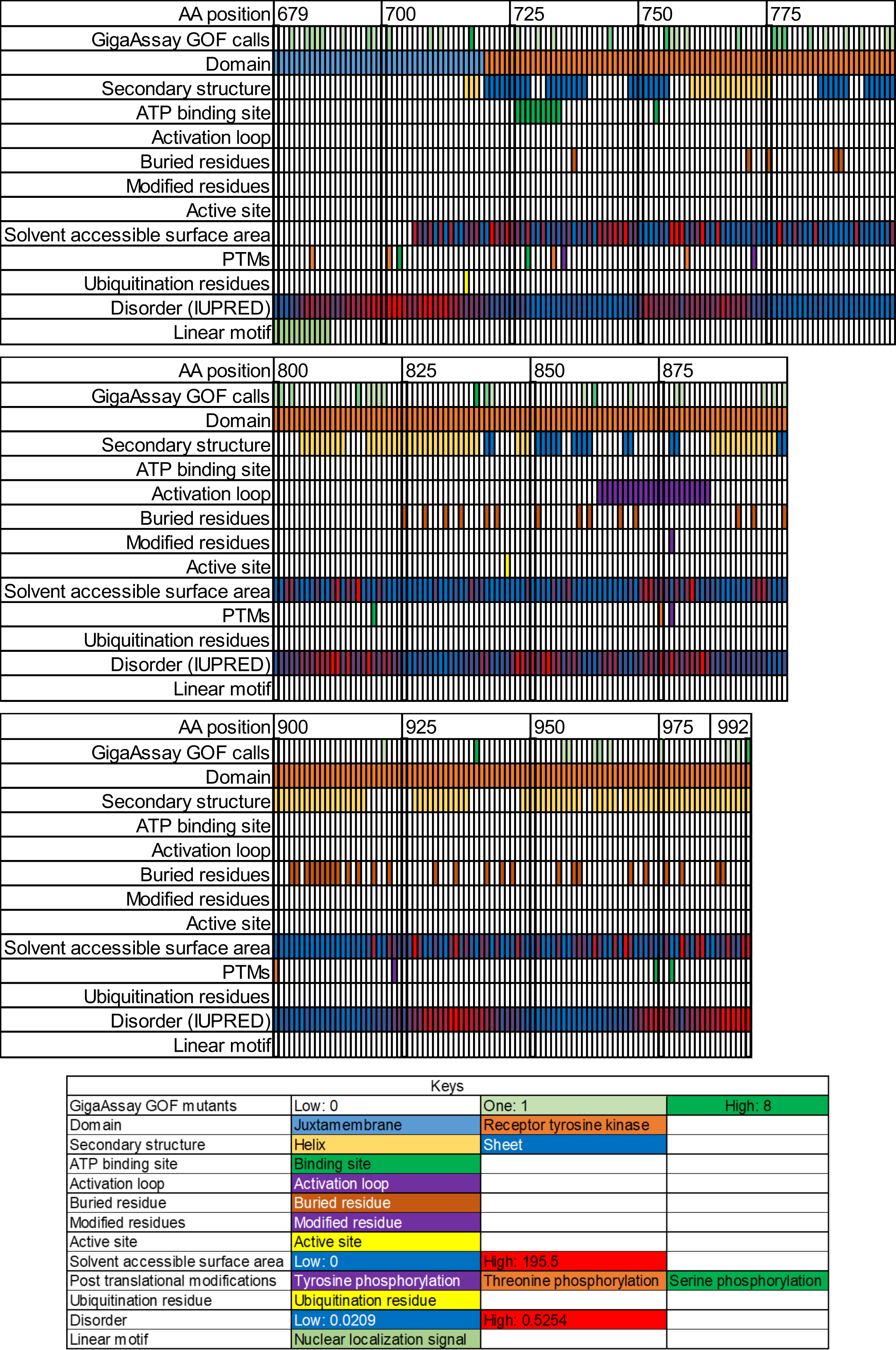
Sequence-function mapping of HER2. Functional domains of the assayed regions (679-705, 708-882, 885-992) are depicted. Each column represents a residue position, while information known about that position are shown in the labeled rows. The number of GOF calls for all possible missense mutants at the position are shown at the top and known functional information about HER2 is shown below. Every 25 amino acids are numbered and have a bolded border.

We examined the spatial relationships and clustering of GOF mutations. For each position, 19 different codons encoding all possible amino acid substitutions were measured in the GigaAssay. Positions with at least one GOF mutation (p <0.05) identified in the GigaAssay were colored on surface and ribbon plots of a published structure of the TK domain of HER2 (PDB: 3PP0: **Fig. 5**); there is no current structure of the conjoined TK and JM domains.[32] GOF mutations in the TK domain are strongly associated with cancer. Several known HER2 oncogenic mutations, including G776S, L755P, L755S, D769H, D769Y, V777L, T798M, V842I, and T862A, are located in the TK domain (**Fig. 5B**). These benchmark GOF mutations are not spatially clustered, but instead were distributed across different regions. In total, 60 GOF positions in the TK domain were identified with the GigaAssay, comprising both novel mutations and previously known ones (**Fig. 5C).**

**Figure 5.**
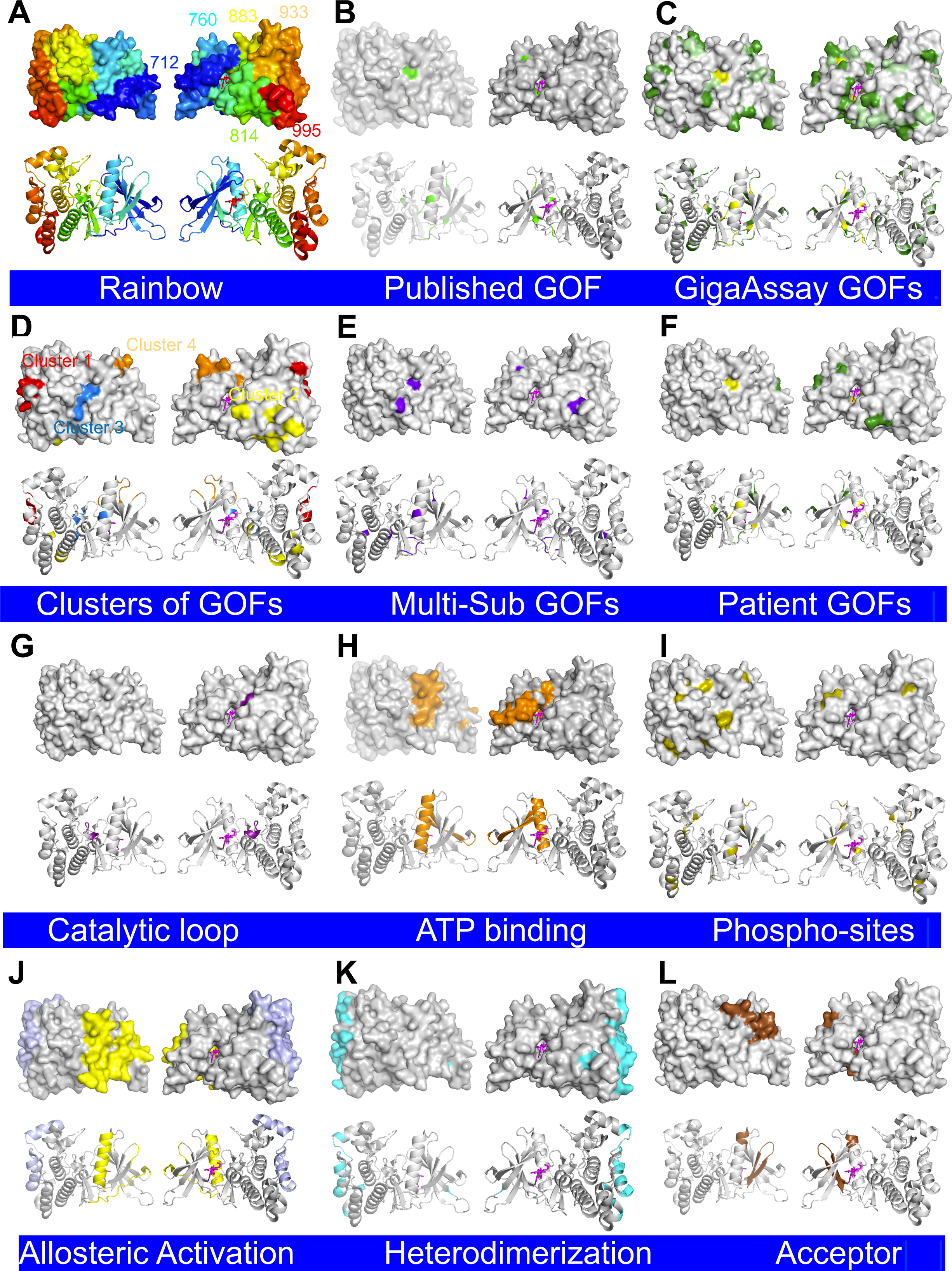
HER2 structure gallery of TK domain structure/function relationships. All surface plots and ribbon diagrams are the WT TK domain HER2 structure (PDB: 3PP0) with one member of each pair rotated 180° about the Y axis: **A.** Position gradient colored as a rainbow. **B.** Published benchmark GOF positions (755, 769, 776, 777, 798, and 862). **C.** GOF positions identified in the GigaAssay, with benchmarks colored yellow, GOF variants requiring a 1 bp codon change are colored pale green, and new GOF positions in the TK domain identified by the GigaAssay colored dark green (726, 730, 733, 744, 755-757, 759, 769, 776-778, 783, 786, 790, 793, 799, 800, 801, 803, 812, 816, 818, 821, 839, 841, 860, 862, 869, 878, 879, 895, 897, 899, 921, 939, 956, 957, 962, 963, 965, 975, 988, 990, and 992). Mutants identified in the GigaAssay that only require a 1 bp codon change (726, 730, 733, 744, 755, 756, 757, 769, 776, 777, 778, 786, 790, 793, 799, 800, 812, 816, 819, 820, 839, 841, 842, 862, 879, 895, 897, 899, 939, 956, 957, 963, 965, 992). **D.** Distinct cluster of GOF mutations (Cluster 1: 939, 956, 957, 962, 963, 965: red; Cluster 2: 812, 818, 819, 820, 821, 988, 990, 992: yellow; Cluster 3: 839, 841, 869, 769, 842: blue; Cluster 4: 730, 755, 756, 757, 759, 793: orange). **E.** Positions with multiple substitutions (Multi-Sub) that are GOF variant positions (769, 755, 992, 776-G778, 783, 839). **F.** Patient GOF variant positions including benchmarks (733, 755, 776, 769, 777, 798, 816, 841). **G.** The catalytic loop (844-850): magenta) [15] **H.** ATP binding site from PDB binding to PubChem ID:03Q SYR127063 ATP mimetic drug (726, 729, 734, 751, 753, 770, 774, 783, 785, 796, 800, 801, 804, 805, 850, 852, 862, 863, 864)[14]. **I.** Phosphorylation (aquamarine, positions 686, 733, 735, 759, 772, 779, 875, 877, 900, 923, 974, 977, 998) **J.** Two allosteric sites in each monomer [yellow (705-718) and light purple (761-765,790-792)] that shift upon the activation dimer. **K.** Heterodimerization with EGFR (706-712, 714, 716, and 718, 761, 764, 765, 768, 772, 790, and another site - 791, 936, 937, 938, 939, 940, 943, 948, 949, 950, 952, 953, 956, 957, 960, 961, 964, 965, 985, 988, 992) [15]. **L.** Acceptor interface for the JM domain (786,790,793,799).

The GOF-centric positions are spatially grouped into 4 distinct clusters (**Fig. 5D**): Cluster 1 contained several new mutations (839, 841, 869) and two previously identified ones (769 and 842). Cluster 2 comprised positions 730, 756, 757, 759, 793, as well as 755 which had been previously characterized. Cluster 3 contained new GOF mutations at positions 939, 956, 957, 962, 963, and 965. Finally, Cluster 4 included GOF mutations at positions 812, 818, 819, 820, 821, 988, 990, and 992. Among the 60 positions, GOF mutations for 36 positions had a substituted codon that required only a 1-base pair (bp) change, making them more likely to occur in cancer patients than 2-bp or 3-bp codon substitutions (**Fig. 5C**, yellow and lighter green).

Of the 60 positions identified, 8 (5 of which were novel) had multiple GOF substitutions with different amino acids. These positions were D769, L755, E992, G776-G778, S783, and V839, with the first three located in the new spatial clusters (**Fig. 5E**). At position D769 larger hydrophobic amino acids (L, W, and F) were associated with GOF activity. Small amino acids at position L755, (G, C, P, S, T, and N) complemented prior knowledge of GOF mutations with GOF small amino acids at this site. Position 776-778 had many GOF substitutions and is consistent with the A775_G776insYVMA indel GOF mutation. Aliphatic amino acids (V, L, M) were favored GOF substitutions at position S783. Basic amino acids (R, H, K, C, N, A) at V839 were particularly enriched in GOF mutations. Hydrophobic amino acids along with several others (F, Y, W, A, M, H) were GOFs at E939. Additionally, five substitutions at E992 (R, K, C, A, and W) were identified as GOF mutations. Two short stretches (positions 706-707 and 883-884) showed complete dropout; therefore, no activity was measured at these sites. Collectively, the density of GOF substitutions at some sites further validates the GigaAssay results.

The GOF mutants at positions 709, 755, 769, 776, 777, 798, and 862, which have been observed in cancer patients did not cluster on the surface with any distinct pattern (**Fig. 5F**). However, some of these patient GOF mutations at positions 755 in cluster 1 and 769 in cluster 2 were next to newly identified GOF positions in these clusters (**Fig. 5D, E**).

We also explored the relationship between GOF mutations in the TK domain and previously reported structural and functional elements. Key determinants within the catalytic loop and ATP binding site, located between the N-terminal and C-terminal lobes, are shown in **Fig. 5G, H**. The newly identified GOF mutations, L726V, A730T, and T733I are located in the phosphate binding loop (positions Leu726–Val734), which is part of the ATP binding site.[14] Similarly, 3 new GOFs (L869K, H878V and A879G) and benchmark variant H878Y are found in the activation loop (positions 863-884), with an additional 4 GOF mutations at position T862, just prior to the loop. This region has been previously noted for its concentration of GOF mutations.[14,33] Notably, T862A is one of the benchmark GOF variants and L869K, which is a chemically similar substitution to L869R, disrupts an autoinhibitory interaction with the C helix and is thought to be a GOF.[8] Although no known functional elements are at positions 776-778, the common activating A775_G776insYVMA indel variant, G776S, a residue that increase HER phosphorylation and signaling, and V777L, a benchmark GOF mutation resides in this region.[14,29,34,35] This region emerged as a major hotspot with 11 new GOF mutations, 2 previously known variants, and a high frequency of variants at positions 776 (57% of possible substitutions) and 778 (63% of possible substitutions had a GOF mutation in at least one replicate). This observation underscores the importance of this region in multiple types of cancer, despite the absence of a known functional determinant.[5,8,14,15,24,36,37] Phosphorylation sites and ubiquitylation sites from the PhosphoSite database are shown in **Fig. 5I** and did not generally overlap with the identified GOF mutations.[38,39]

We mapped several sites that regulate HER2, with insights from EGFR-related activation mechanisms to provide context for understanding GOF mutations.[14] Two sites critical for allosteric rearrangement and activation are shown (**Fig. 5J**).[14] Cluster 3 contains six GOF positions (939, 956, 967, 962, 963, 965) within the allosteric rearrangement regions. Many of the GOF mutations in this cluster correspond to contact residues involved in heterodimerization of HER2 with EGFR (**Fig. 5K**). These findings suggest that disruption of the allosteric rearrangement and/or heterodimerization appears to be strongly linked to GOF activity, although additional experimentation is needed.

Another mechanism linked to GOF mutations is the interaction between the JM and TK domains. The acceptor (**Fig. 5L**) is an 18-amino acid highly conserved interface within the TK domain that binds to the conserved donor region of the JM domain, a relationship also observed in the EGFR structure.[40] GOF mutations in the homologous HER2 regions, both within the acceptor site in the TK domain (786,790,793,799) and the donor site in the JM domain (see next section), include newly identified mutations from the GigaAssay. Notably, position 799 is adjacent to T798M, a gatekeeper mutation for drug resistance.[11] These mutations may disrupt the JM-TK domain intramolecular interaction, potentially leading to the activation of HER2.

### 2.5 Structure - Function of JM domain

The JM domain is a short peptide segment that connects the transmembrane domain to the TK domain in HER2. In many RTKs, the JM domain autoinhibits the kinase activity. The GigaAssay revealed that the JM domain is a hotspot for GOF mutations, with 33% of the 39 positions tested showing at least one mutation with GOF activity. Specifically, GOFs were identified at positions 682, 685-688, 692, 697, 698, 701, 709, 711, and 717 within the JM domain (**Fig. 6**), Positions 706 and 707 were not measured due to dropout in the synthetic variant library.

**Figure 6:**
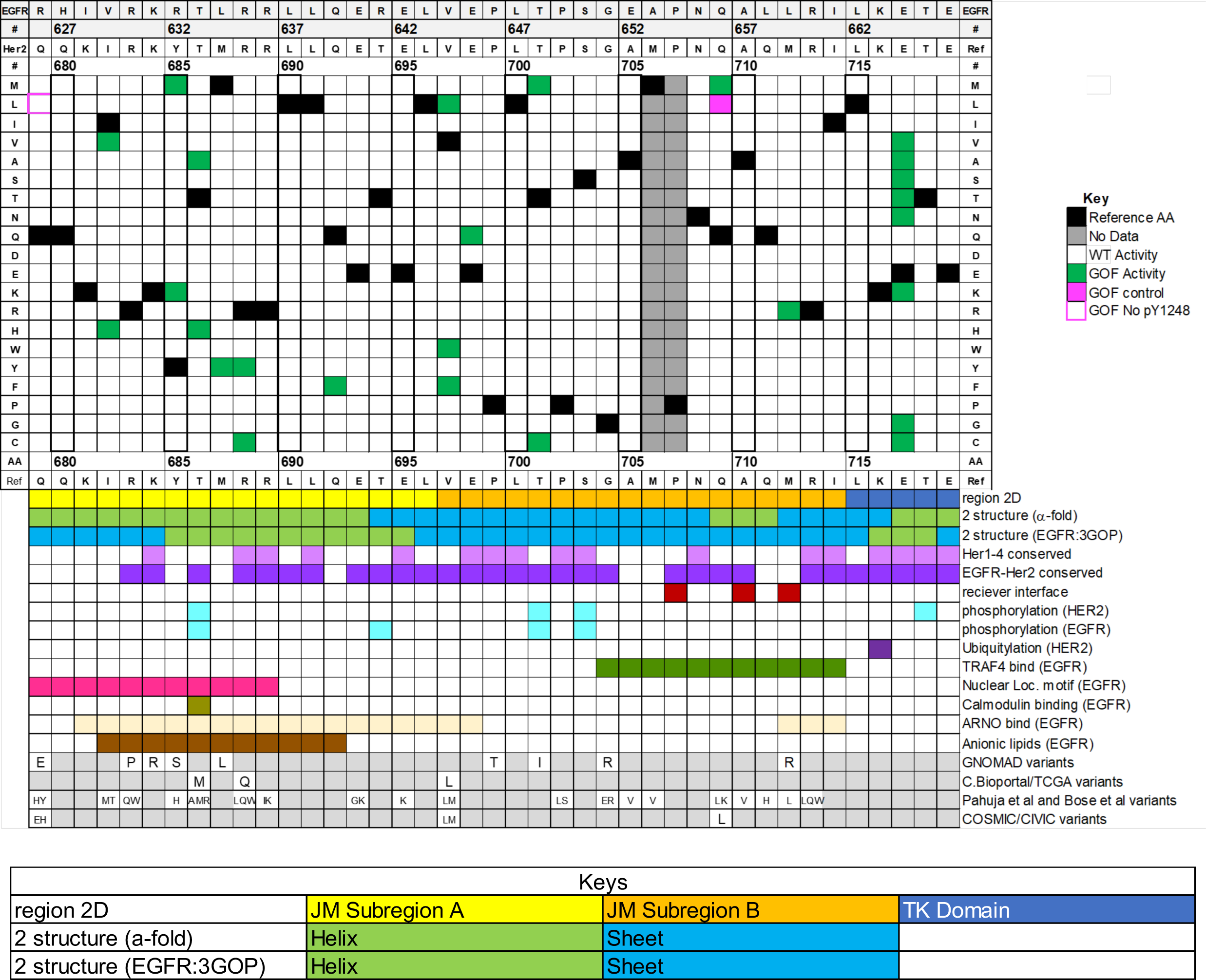
Sequence-function mapping of the JM domain. The GigaAssay results for the JM domain are shown in a heatmap. Labeled functional information is shown in the rows below the activity score heatmap.

A previously reported GOF variant, Q709L was confirmed as a GOF in the GigaAssay and by a flow cytometry assay on a stable cell line (**Fig. 6**, magenta).[14,37] Q679L, another GOF in JM, was previously known to have attenuated phosphorylation, a finding that was confirmed by a flow cytometry assay analysis of a stable cell line.[41] In agreement with these reports, the GigaAssay did not detect significant phosphorylation levels (**Fig. 6**, magenta border).

Much more is known about EGFR than HER2, so we explored if the JM region had high sequence identity among HER family members potentially enabling us to infer functional elements in HER2. When the JM amino acid sequences of HER1(EGFR), HER2, HER3, and HER4 are aligned, 41% of the amino acids are conserved among all family members, indicating high degree of conservation of this segment (**Fig. 6**). A pairwise alignment of EGFR with HER2 shows even greater conservation at 73%. Both EGFR and HER2 are phosphorylated at two positions, 686 and 701 in HER2, which both correspond to newly identified GOF mutations from the GigaAssay. T686A and T686H, may play a similar role to three known substitutions at position 686 that are also found in tumors, suggesting an important role for this phosphorylation site, although this residue is also in a binding site for calmodulin.[14,42] There are 3 additional phosphorylation sites in this region that do not have any GOF mutations.

The JM domain spanning positions 679-697 binds various functional elements in the analogous regions of EGFR. This includes a nuclear localization sequence (NLS), calmodulin, ARNO, and anion lipids. The same region in HER2 has at least one GOF at each of these binding sites, marking it as a hotspot of GOF mutations with 14 newly identified GOF mutations. Additionally, the Traf4 binding site, which contains the previously known GOF, Q709L, also has two new GOF mutations, Q709M and Q711R at this site.[43] In the analogous 3D structure of EGFR, part of the JM domain interacts with the receiver interface in the TK domain, which activates EGFR. We tested two of the three positions in this region and identified Q711R as a GOF mutation within this functional element.

Among the positions with multiple GOF substitutions, E717 stands out with 8 of the 19 potential substitutions being GOF mutations. E717 is positioned adjacent to K716, a residue known to be ubiquitylated (PhosphoSite).[38] One possible explanation of the observed GOF activity is that a substitution at E717 may disrupt ubiquitin-mediated degradation, leading to persistent GOF activity. In summary, our findings identify several new GOF mutations linked to key functional elements in the JM domain, further highlighting its role in HER2 activation.

Structurally, no existing model of the conjoined JM and TK domains of HER2 was available, so we predicted one using AlphaFold V2.0.[32] For comparison, we examined a structure of EGFR that contains both the JM and TK domains that share high sequence identity and similarity. The two structures showed overlap in their secondary structure elements predominately comprising α-helices and random coils. Notably, a large α-helix, which is particularly rich in GOF mutations contains binding sites for several functional elements including binding sites for the NLS, calmodulin, ARNO, anion lipids), Additionally, a smaller α-helix overlaps the 8 new GOF mutations located at E717.

We investigated structure-function relationships within the JM. Some of the functional elements were inferred by strong sequence identity with EGFR in this region. Mutants and functional elements in the JM region were mapped onto a predicted structure generated using AlphaFold V2.0.[32] For each surface map, both ribbon and surface representations are provided, with one pair member rotated 180° about the Z axis. A rainbow-colored surface plot shows the approximate location of numbered positions (**Fig. 7A**). When the autoinhibitory fragment that interacts with the TK domain shown in **Fig. 7B** is compared to the *in vitro* GOF mutations shown in **Fig. 7C**, several mutations overlap. This suggests that certain GOF mutations (V697F/W/L, E698Q, T701M/C) may relieve JM-mediated inhibition of the TK domain.[5,14] **Fig. 7D, E** show the strong sequence conservation of the JM domains in HER2 and EGFR, as well as across all HER family members including the conserved positions corresponding to the three suspected autoinhibitory GOF residues.

**Figure 7.**
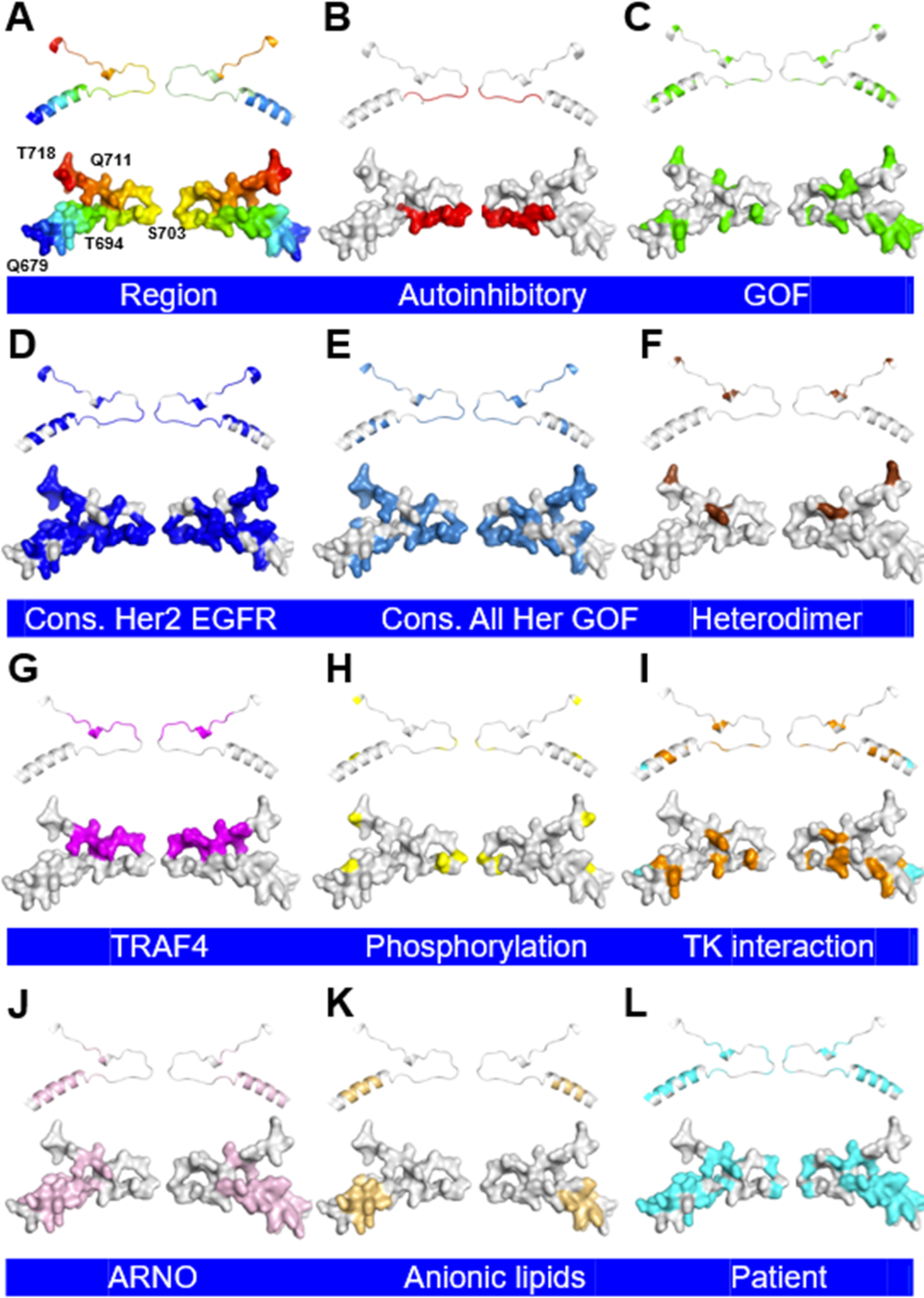
HER2 structure gallery of JM domain structure/function relationships. All surface plots and ribbon diagrams are of the predicted WT HER2 structure of the JM domain with AlphaFold.[32] One member of each 3D structure pair rotated 180° about the Y axis: **A**. Position gradient colored as a rainbow; **B**. Conserved autoinhibitory segment of the JM region in EGFR; **C**. Positions with GOF variants identified in the GigaAssay; **D**. Conserved residues with EGFR; **E**. Conserved residues with all HER family members; **F**. Heterodimerization positions of the JM region with EGFR; **G.** Positions that bind to TRAF4 in EGFR; **H.** Positions in HER2 that are phosphorylated (PhosphoSite); **I.** Conserved autoinhibitory residues that bind to the TK domain in the JM region in EGFR [55]; Benchmark mutation 679L is shown in cyan; **J.** Positions that bind to ARNO in EGFR; **K.** Positions that bind to Anionic lipids in EGFR; **L.** Positions with variants observed in cancer patients. A ubiquitylation site is at position 716 (not shown).

Another potential mechanism of JM domain-mediated autoactivation of the TK activity is through heterodimerization with EGFR.[14,44] In the homologous region in HER2, a previously known GOF mutation, Q709L GOF is positioned near several newly identified GOF mutations including Q709M, Q711R, and E717G/A/C/S/T/N/V/K, all of which are within this dimerization interface (**Fig. 7F**).[14]

Additionally, the JM domain contains several key protein and lipid interaction sites, including an anionic lipid binding site, a nuclear localization sequence, and an ARNO binding site. These sites are in close proximity with the TK domain and portions are conserved and enriched with a cluster of GOF mutations (compare **Fig. 7F, I, J** and **L** to **D** and **E**).The TRAF4 binding site, which contacts the autoinhibitory segment, may also activate the TK based upon its known effect on EGFR (**Fig. 7G**).[45] This region also shows significant overlap with oncogenic variants identified in patients **Fig. 7L**. The clustering of GOF mutations at functional sites on the surface map further underscores the sensitivity of the JM domain in HER2 activation.

### 2.6 Analysis of Patients with HER2 GOF mutants

HER2 somatic missense mutations are associated with poor survival outcomes.[5] In our GigaAssay, we identified 112 novel GOF variants which were consistently reproduced across technical replicates. To assess their clinical relevance, we sought to determine how many of these variants had been observed in human tumor specimens. To do this, we compiled data from multiple sources including the TCGA, ClinVar, COSMIC.[46–48] These data sets together contain information on 15,000 tumors specimens, including several thousand with a *ERBB2* (HER2) missense mutation. While this dataset is enriched for breast, lung, and colorectal neoplasms, it is not limited to any specific type of tumor.

From these samples, 109 unique variants were identified, 19 of which were identified as GOF mutations in the GigaAssay experiment (**Fig. 8A**). Of these, 10 were the previously mentioned benchmark variants, which were not only confirmed in the GigaAssay, but also detected in tumor samples. In total, the GigaAssay identified 112 new GOF variants, of which 9 were found in patient tumors (V697L, E717K, E171A, T733I, G776V, V777M, L841V, L755A, E812K). A vast majority of novel GOFs identified in the GigaAssay are rare or even nonexistent; however, in the future they may be found in patients’ tumors.

**Figure 8:**
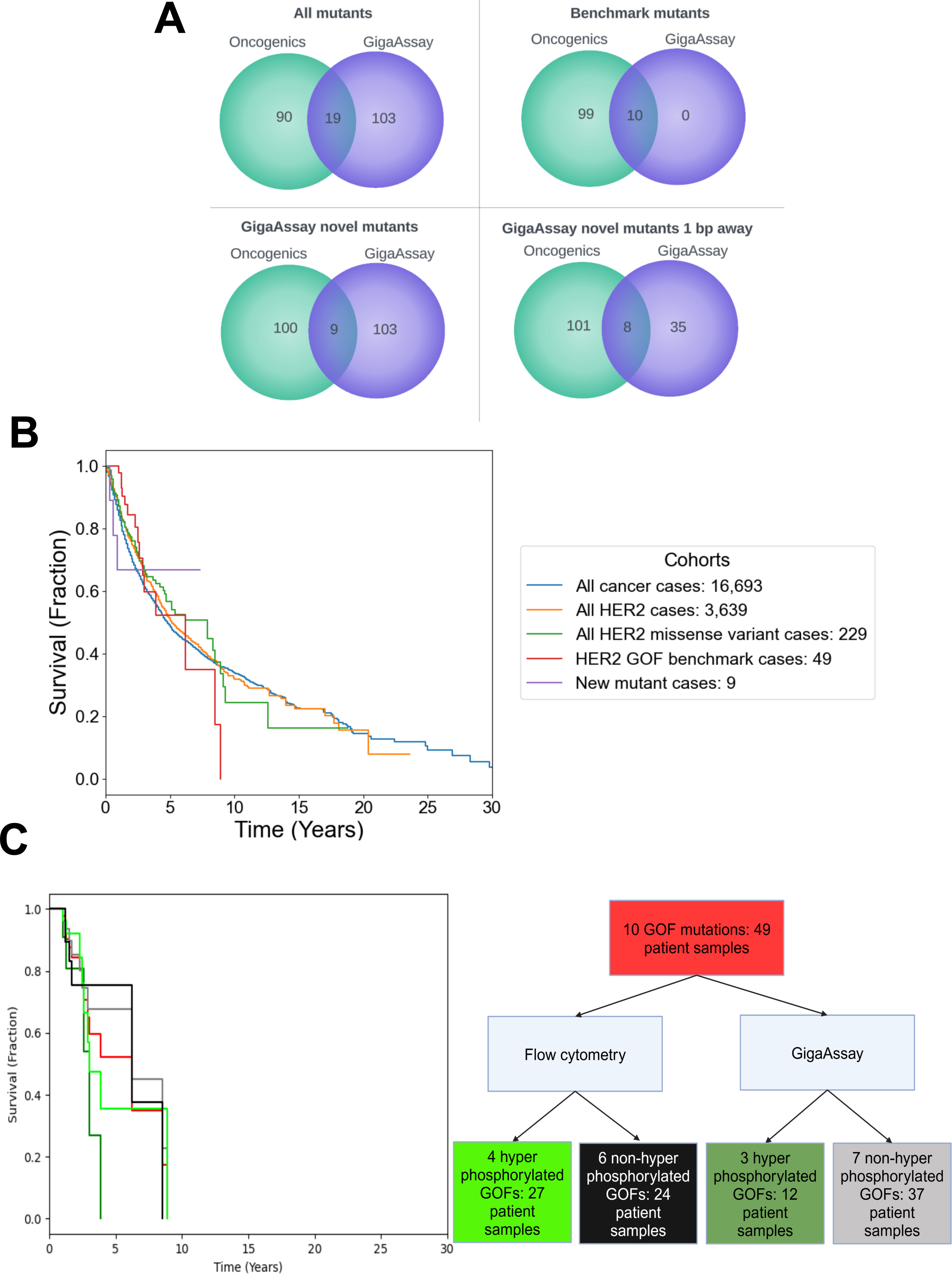
Comparing HER2 mutants in cancer patient tumors to GigaAssay GOF variants. Tumor mutant data was obtained from individual publications, NIH GCD, ClinVar, COSMIC, and The Cancer Genome Atlas. 109 mutants from these data sources were compared to various sets of mutants with GOF calls from the GigaAssay. **A.** For each category, number of mutants found amongst the tumor sources (green) and number of mutants with GOF calls from the GigaAssay (Blue). **B, C.** Kaplan Meier plots representing survival of patients with various categories of mutations. These cohorts contain variant information known prior to and after the GigaAssay experiments. **B.** Survival of various cohorts of cancer patients with different sets of genetic dispositions. The first four cohorts are groups defined by prior knowledge of HER2 mutant status and survival; the last group consists of patient samples containing at least one novel GigaAssay GOF mutant. **C.** Comparison of survival of patients with tumors with different sets of GOF benchmark mutations. GOF benchmarks with visual indication of hyperphosphorylation relative to the other benchmarks are separated into groups. The groups are defined by the experiment in which the hyperphosphorylation is observed. Patients with GigaAssay hyperphosphorylation GOF variants have poorer survival.

Many GOF variants identified with the GigaAssay encode amino acid substitutions that require 2- or 3-bp changes to create the codon for the substitution. These mutants are far less likely to coexist within the same tumor, but may still exhibit oncogenic potential. Both sets of mutants have utility: the total set of GOF mutants (n = 122) provides insight into the structure-function relationship and oncogenic potential of each position and substitution, while the mutations possible with a single nucleotide substitution (n = 51) are more likely to occur in tumors and be oncogenic.

Among the 10 benchmark variants, 8 are possible with a 1-bp substitution. However, L755P and T798M, while considered oncogenic mutations in the literature, require a 2-bp substitution.[8,12,15,23] This observation is supported from an analysis of tumors from 16,693 patients, where these mutations were not observed. We were surprised that 35 of the 1-bp GOF variant positions in the GigaAssay did not occur in any of the tumor specimens. The individual mutants and their occurrence in tumors are detailed in **Table 1**. Additional information for the 1-bp substitutions with GOF activity can be found in **Supplementary Fig. S9.**

**Table 1.** Data sources of oncology-associated variants.

| Mutant | Control/Novel (C/N) | Individual Sources | NIH GDC | Clinvar | COSMIC | CKB | TCGA |
| --- | --- | --- | --- | --- | --- | --- | --- |
| Q709L | C | X |  |  |  |  |  |
| L755S | C | X | X | X | X |  | X |
| L755P | C | X |  | X |  |  |  |
| D769Y | C | X | X | X | X |  | X |
| D769H | C | X | X | X | X |  | X |
| G776S | C | X | X | X |  |  |  |
| V777L | C | X | X | X | X |  | X |
| T798M | C | X |  |  |  |  |  |
| T862A | C | X | X |  | X |  | X |
| V842I | C | X | X |  | X |  | X |
| V697L | N | X | X |  | X | X | X |
| E717K | N | X |  |  | X | X |  |
| E717A | N | X |  |  |  |  |  |
| T733I | N | X | X |  |  |  | X |
| G776V | N | X | X |  | X |  | X |
| V777M | N | X | X | X |  | X | X |
| L841V | N |  |  |  | X |  |  |
| L755A | N |  |  |  |  | X |  |
| E812K | N |  |  |  |  | X |  |

Since we identified a benchmark set of mutants validated by flow cytometry and the GigaAssay, as well as new GOF mutations that occurred in tumors, we wanted to further understand how these mutants affect patient survival. The GDC database has genomic data from tumor biopsies, as well as associated patient survival data (n = 16,693 cases).[49] We searched the database for tumors harboring *ERBB2* variants (n = 3,639 cases) in any type of tumor. We compared the datasets with a custom script tool to create Kaplan-Meier plots. We did not notice any difference in survival for tumors that had *ERBB2* missense variants when compared to total cases, in which only 20% of patients survived after 17 years (**Fig. 8B**).

We next assessed the benchmark *ERBB2* GOF variants and identified 49 cases across the 10 benchmark mutants. Among these, only 20% of the patients survived beyond 8.5 years, indicating a more vulnerable population. In contrast, when we examined the newly identified mutants from the GigaAssay, we found only 9 patients. Notably, only 20% of these patients survived after 2.7 years, suggesting that these GOF variants are even more deleterious, though they are rare. This small set of mutants contributed to a 16% increase in diagnostic yield. Additionally, these mutants were also found across multiple patient data sources (**Table 1**).

Since all *ERBB2* GOF variant were analyzed under identical GigaAssay conditions, we investigated whether stronger activation of HER2 correlated with poorer patient survival. First, we assessed the relative activation states of previously characterized HER2 GOF mutants using flow cytometry. The L755P, L755S, V777L and T862A mutants exhibited significantly higher hyperphosphorylation levels compared to other GOF mutations.[22] Similarly, the V777L, T862A, and T798M variants displayed greater hyperphosphorylated levels than the benchmark HER2 GOF mutants in UMI-barcode activity profiles from the GigaAssay experiment.[22] When survival data for patients with these GigaAssay variants were analyzed separately, their patient survival was halved when compared to that of patients with well-known HER2 GOF variants (**Fig. 8C**).

A subset of the mutants demonstrated elevated hyperphosphorylation in both a flow cytometry assay (n = 4) and the GigaAssay (n = 3) appeared in 27 patients. The only discrepancy between the two methods was the L755S mutant, which was detected by flow cytometry, but not by the GigaAssay. Additionally, the L755P and T798M mutants were absent in patients from this cohort across both methods. Patients with V777L and T862A mutants, which exhibited strong hyperphosphorylation in the GigaAssay, demonstrated a two-fold worse survival outcome, as expected (**Fig. 8C**). In contrast, adding the L755S mutant to this set, all showing hyperphosphorylation by flow cytometry, had survival rates similar to those of other benchmark mutants. While the benchmark data supported a quantitative assessment of hyperphosphorylation levels, this was not observed for the GOF mutants in the UMI-barcode activity profiles from the GigaAssay. The GigaAssay activity score showed a weak negative correlation (R^2^ = 0.11) with survival, suggesting that these GOF variants may be associated with poor patient outcome (technical replicates were averaged; **Supplementary Fig. S10**).

## 3. Discussion and Conclusions

*ERBB2 (*HER2) oncogenic mutations have been studied for over two decades, following the discovery of somatic *ERBB2* mutations in the TK domain of primary lung tumors.[50] From the literature, we selected 10 well-characterized missense GOF mutations of HER2, which are linked to hyperphosphorylation. These GOF mutations have been extensively investigated by multiple research groups, each using various combinations of techniques to identify the effect of causal driver mutations and the association of these variants with different tumor types. A typical workflow consists of testing the effect of such variants individually in the cancer cell type of interest. However, this non-uniform workflow may introduce errors and artifacts and result in misinterpretation of the effect of cancer variants. Therefore, a standardized workflow is needed to better assess and compare the relative impacts of different oncogenic mutations. Our study, too, highlights the need for such standardization in the analysis of *ERBB2* GOF variants.

We identified 6 aspects of this study that are unique:

1. **Discovery of New GOF Mutations**: We identified 112 novel, *in vitro* HER2 GOF mutations with the GigaAssay, including 9 that have been identified in cancer patients in publicly available data sources, but had not been previously highlighted.
2. ***In vitro* vs. Patient Samples**: The presence, or lack thereof for many *in vitro Erbb2* GOF mutations in cancer patient samples suggests that an insufficient number of patient biopsies have been analyzed.
3. **Mechanistic Insights**: Five spatial clusters in the TK and JM domains are enriched with GOF mutations, supporting GOF activation by two major mechanisms: (1) allosteric rearrangement and heterodimerization with EGFR, and (2) binding of the JM domain to the TK domain (**Fig. 9**).
4. **Alternative Mechanism for Cancer**: Analysis of the L726F, H878Y, and Q679L mutations, both from previous reports and in this study, suggests a secondary, less prevalent, Y1248-phosphorylation-independent mechanistic pathway driving cancer.
5. **Enhanced Diagnostic Yield**: New GOF variants increased diagnostic yield by 20% in a small cancer patient cohort.
6. **Prognostic Implications**: New GOF variants predict better patient survival than those with previously known GOF variants. This may be because the most severe variants are more commonly found in patients to this point.

**Figure 9:**
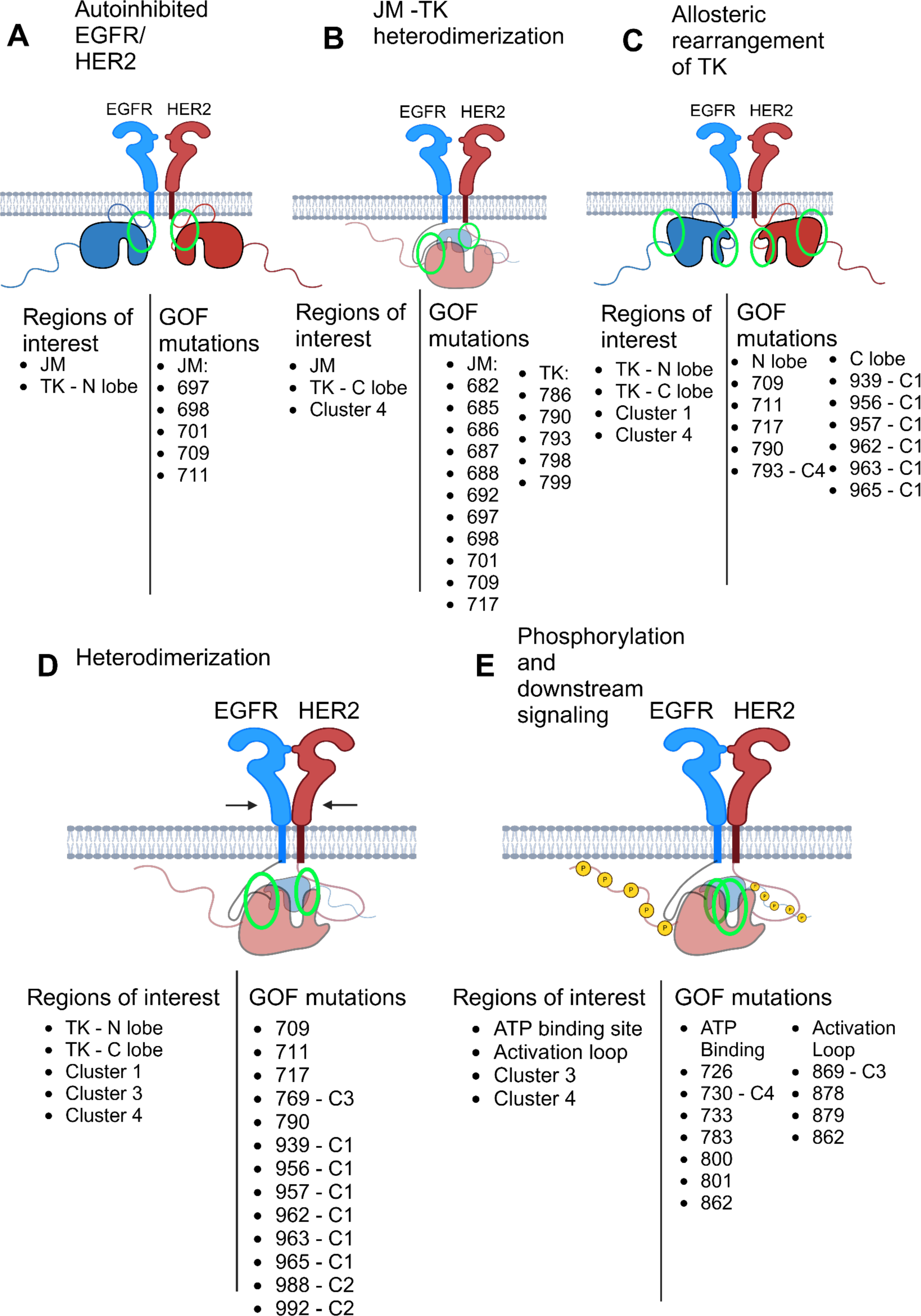
Overview of different mechanistic steps activating HER2. A-E. Steps in the HER2 activation pathway are shown as a schematic representation, listing regions and previously characterized GOF mutations and newly identified GOF mutations for each step. The cluster location of each GOF is indicated (C1-C4). **B, D, E**. Some models have a smaller EGFR TK domain reflecting its position behind the HER2 TK domain.

### 3.1 HER2 GigaAssay performance

In this study, we performed saturation mutagenesis of all amino substitutions in the TK (residues 715-992) and JM (residues 679-719) domains of HER2 using the GigaAssay. These regions were selected due to their relatively high frequency of somatic mutations in cancer specimens. The *ERBB2* plasmid library for the GigaAssay contained over 99% of the expected 5,966 mutants, with the few missing variants originating from 4 specific positions and likely a result of synthesis dropout. The pHER2 GigaAssay results were then filtered to exclude variants with low read counts, less than 2 UMI-barcodes, and those identified as arising from common errors in oligonucleotide synthesis. The filtered final datasets for each replicate included 5,886 variants, representing 99% of the intended mutants (**Supplementary Files 1 & 2**).

In the Giacoletto et al, the GOF activities and classifications of GigaAssay mutants were compared to a benchmark literature dataset derived from the literature identifying 10 well-characterized GOF mutants, and 7 non-GOF mutants with WT levels of activity.[22] The 100% statistical accuracy observed in this experiment represents an improvement over the 94% accuracy of our previous GigaAssay using HIV Tat.[13] This improvement may reflect advancements in experimental protocols and bioinformatics since the initial publication of the GigaAssay, or it may simply reflect variability between experiments.[13]

In a recent paper, we proposed that scientific methods should aim to become a High Accuracy and Throughput (HAT) assay with >99% accuracy and a throughput of at least 1,000 measurements.[52] Thus far, the only established HAT assays are based on NGS, which meets these criteria. The success of the HER2 GOF GigaAssay suggests that the GigaAssay experiments reported here support its designation as the next HAT assay; however, further validation across multiple GigaAssays for different genes is needed. The integration of HAT Assays with advanced machine learning approaches is expected to usher in a new era of scientific discovery, which we have termed the HAT Era.[52]

### 3.2 Discovery of new GOF mutations

The first *ERBB2* (HER2) driver mutation was discovered in 2006 and since then, several oncogenic mutations across the extracellular, JM, and TK domains have been characterized in the scientific literature. In our study, we focused on 10 well-characterized missense GOF mutations as benchmark GOF controls. As anticipated, we validated both GOF and 7 non-GOF control variants in our flow cytometry assay for pHER2 and the same mutations validated in our GigaAssay. Surprisingly, the control GOF mutations do not cluster together, either along the primary sequence or within regions of the 3D structure of the HER2 TK domain (**Fig. 5**).

The GigaAssay not only confirmed the activity of the known control mutations, but also uncovered 112 new GOF mutations. This is a striking increase over previously identified GOF variants, which is particularly surprising given the extensive research on HER2 in cancer. Several key observations support the validity of these new GOF mutations: 1) The high accuracy of the GigaAssay with controls; 2) The new GOF mutations cluster at the same or adjacent positions in the primary sequence; 3) The new GOF mutations spatially cluster within specific regions of the HER2 3D structure; 4) Many of the mutations are located in critical regions related to HER2 activation, including the EGFR heterodimerization site and in the regions undergoing allosteric changes upon TK activation[14,15]; and 5) Several of these GOF variants also present as somatic variants in previous sequence analysis of cancer biopsies.

Although the HER2 GOF control mutants do not cluster in the sequence or structure, distinct patterns were observed for the newly identified GOF mutants. There were 4 distinct clusters found among the newly identified GOF mutants (**Figs. 5** and **7**). Cluster 3, in particular, includes 19 new GOF mutants, situated within or adjacent to the HER2 heterodimerization site with EGFR (936-940, 943, 948-950, 952-953, 956-957, 960, 961, 964, 965, 985, 988, and 992), as well as other HER family members.[14]

Another region of the EGFR interaction site in HER2 is in the JM domain. Two previously identified variants in this region of HER2 are Q679L and Q709L.[4,14,37,53] The GigaAssay revealed a 5^th^ cluster of GOF mutations at positions 682, 685-688, 692, 697, 698, 701, 709, 712 and 717. Notably, some of these GOF positions (709, 711, 716, and 718) are located within or adjacent to the EGFR-HER2 heterodimer interface, where they bind to the acceptor site in the TK domain.[14] These findings further reinforce a role for this mechanism in oncogenesis.

The JM domain has many additional GOF positions, suggesting that other activation mechanisms are also at play. This region autoinhibits kinase activity, a property shared with other RTKs. For example, V560G in KIT and V561D in PDGFRA are known to drive oncogenic signaling.[54] The GigaAssay identified a cluster with 12 of the 32 positions within the HER2 JM domain, supporting its role in autoinhibition of the kinase activity.

While the structural basis for the EGFR JM domain inhibition has been established, the corresponding structures for HER2 do not contain both domains (e.g. PDB id: 3GOP).[55] Given the 74% sequence identity between the JM domains of HER2 and EGFR, the known autoinhibitory nature of the HER2 JM domain, we assessed if the GigaAssay findings support extrapolation of the EGFR mechanism to HER2.[31,55] In EGFR, the region corresponding to positions 696-712 in HER2, known as the “latch” or “receiver”, is the determinant for binding the “acceptor” in the TK domain interface. For HER2, residues 697, 698, 701, 709, and 711 in the acceptor region are new GOF positions and include Q709L, a previously known GOF, are involved in the TK activation. [4,14,31,37] Notably, conserved residues in this region in EGFR bind TRAF4, a RING finger protein that activates the kinase.[45]

The EGFR structure features an α-helix in the JM domain from residues 686-695 that does not interact with its kinase domain.[55] Similarly, the HER2 JM domain contains GOF mutants in this region, specifically at positions 682, 685-688, and 692, suggesting that this α-helix also contributes to kinase activation in HER2 (**Figs. 6** & **7**). In EGFR, this region binds to anionic lipids, ARNO, calmodulin.[56] Given the sequence between the JM domains of HER2 and EGFR, these factors may also play a role in activating the HER2 kinase by binding to the corresponding amino acids in HER2.

In conclusion, the new 112 GigaAssay GOF variants have provided additional coverage over the 10 previously characterized GOF variants for HER2 and provide a new context to interpret these mutants to better understand the most critical molecular function and oncogenic mechanisms.

### 3.4 GOF variants associated with cancer data

A major question arising from the GigaAssay experiment is why so many novel HER2 GOF variants (n =112 variants) were identified, despite not being previously associated with cancer in patients. In an analysis of approximately 3,700 tumor samples with HER2 mutations, only 9 new GOF variants were found as somatic variants in patients. One possibility is that not enough tumors have been sequenced yet. However, there are several other explanations that can account for this discrepancy.

First, nearly half of the GOF variants identified with the GigaAssay encode amino acid substitutions that require 2- or 3-bp changes (n = 71), making them less likely to occur in patients. However, these mutations provide valuable insights into HER2 structure / function relationships, help identify or validate causal oncogenic mechanisms, and may emerge as variants of uncertain significance (VUS)s in future patients. Additionally, rare multi-bp changes may occur in patients. For example, both the L755P (WT codon = TTG) mutant and the novel L755A variant requires at least a 2-bp change and these are the only 2-bp change mutants observed in patients. Interestingly, these variants occur at the same position as the L755S mutant (which requires only a 1-bp change), the most common HER2 variant in cancer patients.[12,57]

Given a cellular mutation rate of 2.2 ×10^−9^ per bp per year, 2-bp codon mutations would be extremely rare, suggesting they are unlikely to occur in a human lifetime, even if all individuals lived to 100 years.[58] One potential explanation is that mutations in specific dinucleotides, such as pyrimidines (as observed in p53) are more favorable; however, this trend was not observed for these variants.[59] A more likely plausible explanation is that these 2-bp mutations arise through sequent changes. For instance, the WT L755 (TTG) reference codon could undergo a 1-bp change to produce the oncogenic L755S (TCG) mutation. A subsequent mutation of this codon could give rise to a L755A (codon = GCG) or L755P (codon = CCG), both of which require a 2-bp change. The T798M, although a GOF variant, is typically associated with acquired resistance, supporting the idea of a multiple step process.[60] While sequential mutations might explain the appearance of the 2-bp codon changes, we cannot rule out that an unknown mechanism selectively favors these very rare variants.

An unknown mechanism may negatively select against many 1-bp codon GOF variants that could otherwise occur in patients. Nonetheless, the GigaAssay identified 35 new GOF variants that only require 1-bp changes for amino acid substitutions and did not occur in patients. Interestingly, these variants were not found in major databases or reported in the literature. Notably, these mutations clustered in both sequence and structural regions alongside those positions that require 2- or 3-bp changes per codon, further supporting their potential significance (compare **Fig. 5E, F**).

Given the high accuracy and comprehensive nature of this saturating functional analysis, our study provides a rare opportunity to better understand tumor mutational signatures and infer potential mechanisms. There are several possible explanations for the observation of the 35 new GOF variants in the GigaAssay, but not in patients.

These mutations may be absent in patient tumor biopsies for several reasons: 1) they never occur; 2) they occur, but are subsequently removed; or 3) phosphorylation at this site may not contribute to tumorigenesis. While the models for variability of the reference codons, their substitutions, and their interpretation are highly complex and beyond the scope of this paper, we discuss a few potential mechanisms:

1. **GOF variants may not appear** due to factors such as limited chromatin accessibility, which may hinder occurrence. Additionally, there could be mutation hotspots driven by codon bias or mutational pathways.[61,62]. However, we did not observe codon bias in GOF variants (data not shown). We also explored whether this subset of GOF variants was enriched in transversion mutations, given that transitions are more common than transversions in the COSMIC database, but found no such enrichment (data not shown).[46,62]. Another possibility is that DNA repair is more efficient at these positions, as previously suggested.[63] Physiological factors such as selective apoptosis or other forms of cell death, may also play a role. At least some of mutations may be benign or lower penetrance *in vivo*, so are never realized since the patients don’t develop a detectable tumor. Or, it could be simply be that an insufficient number of tumors has been sequenced or analyzed.
2. **GOF variants that do appear** may be removed by various DNA repair mechanisms. There are 26 known mutational signatures that could influence the repair efficiency of GOF variants, potentially preventing their persistence.[63]
3. **Phosphorylation may not be oncogenic** in some cases, as certain variants may be phosphorylated at Y1248, but fail to activate HER2 or induce cancer. It’s possible that mutations at these positions make the protein more susceptible to dephosphorylation by phosphatases, or they may induce interactions with other proteins that do not suppress tumorigenesis.

### 3.5 Y1248-phosphorylation independent mechanistic pathway

Although the Y1248 phosphorylation site is often considered a marker of HER2 activation, it is not essential for signaling to the Erk and Akt pathways.[64] Several reported oncogenic mutants have low or no pHER2 [23,24,41,65]. For example, HPAFII cells expressing Q679L exhibit no increase in pHER2, while its expression in HPNE cells leads to elevated pHER2 levels, suggesting cell line-specific differences in the autophosphorylation of this mutant.[26] Similarly, expression of H878Y in HEK293, BaF3, and BEAS-2B cells results in only a modest increase in pHER2 compared to cells expressing the WT protein.[26] Finally, MCF10A cells expressing L726F display reduced phosphorylation at multiple sites compared to those expressing WT protein.[65] These reduced phosphorylation levels were confirmed by flow cytometry and in the GigaAssay.[22]

One possible explanation is that these mutants may better recruit a protein tyrosine phosphatase, a mechanism that has been implicated in the hypophosphorylation at L726F.[23] Taken together, these findings suggest that these mutants are oncogenic through a Y1248-phosphorylation independent pathway, and further research is needed to elucidate the precise mechanism involved.

## 4. Methods

### 4.1. Molecular Biology

#### Generation of UMI-barcoded variant plasmid libraries

A doxycycline-inducible lentiviral plasmid was made by PCR amplifying the TreTIGHT doxycycline-inducible promoter from pSSI9343 (a kind gift from Sierra Sciences, Reno, NV) subcloned into pLJM1_MCS. We engineered a lentiviral plasmid containing the doxycycline-inducible TreTIGHT inducible promoter with doxycycline. The TreTIGHT doxycycline-inducible promoter was obtained via PCR amplification (a kind gift from Sierra Sciences in Reno, NV).

A double-stranded (ds) DNA library containing codon-optimized full-length *ERBB2* cDNAs with sequences for all the possible single amino acid mutants in the JM and TK domains was synthesized by Twist Bioscience (San Francisco, CA). The library cDNAs were subcloned into the lentiviral plasmid and 32 nucleotides of randomized UMI-barcode was added into the 3’ untranslated region (UTR) by extension PCR. The purified and dialyzed assembly reaction mixture was electroporated into Endura electrocompetent cells (Lucigen, Middleton, WI). Transformants were scraped, plasmid DNA was isolated, and the full-length *ERBB2* cDNA and UMI-barcode regions of the GML plasmid library was sequenced with PacBio at the DNA Sequencing Center, Brigham Young University, Salt Lake City, UT. PCR (15 cycles) was used to generate DNA fragments for long-read sequencing using Q5 High Fidelity DNA Polymerase (New England Biolabs, Ipswich, PA).

#### Construction of individual *ERBB2 variant alleles*

pcDNA3.1+/C-(K)-DYK-ERBB2 containing the full-length WT *ERBB2* cDNA from pcDNA3.1+/C-(K)-DYK-ERBB2 (NM_004448; NP_004439; GenScript, Piscataway, NJ) was PCR amplified and subcloned into the pLJM1_MCS lentiviral plasmid. The A775_G776insYVMA *ERBB2* GOF control mutant was generated by PCR mutagenesis and subcloned into lentiviral plasmid pLJM1_MCS.[29] All other HER2 missense mutation controls were made using QuikChange Lightning Site-Directed Mutagenesis Kit (Agilent, Santa Clara, CA) and PCR mutagenesis. Mutant *ERBB2* cDNA was subcloned into a lentiviral expression vector.

#### Deep sequencing of UMI-barcoded variant libraries

For deep sequencing, PCR primers flanking the UMI-barcode region were used for targeted deep sequencing of the *ERBB2* cDNA library. Staggered nucleotides were added via PCR primers between the Nextera Read Adaptor sequences and targeted UMI region to increase library diversity for Illumina sequencing. gDNA extracted from sorted cell populations was split and PCR amplified for 25 cycles in multiple reactions. A size-selected PCR product from the first PCR (purified using SPRIselect beads) was amplified by PCR to add dual-indexes and the P5 and P7 sequences on 5’ and 3’ ends, respectively. PCR was performed for 6 or 8 cycles, PCR products ranging were purified by size exclusion with SPRIselect Beads (Beckman, Brea, CA). NGS libraries were quantified using a Qubit dsDNA HS Assay Kit (Thermo Fisher Scientific, Hampton, NH) and checked for the expected size using the DNA 7500 Kit for the Bioanalyzer 2100 (Agilent, #Santa Clara, CA). Samples were sequenced on either a NextSeq 500 in the Genomics Acquisition and Analysis Core at the University of Nevada, Las Vegas or on a NovaSeq at CD Genomics (Shirley, NY).

### 4.2 Cell culture

HEK-293 cells expressing TET-response element were used to generate cell lines for all experiments. Cells were cultured in Dulbecco’s Modified Eagle medium (DMEM) media. Stable cell lines expressing HER2 control variants or GML plasmid libraries were transduced with recombinant lentiviruses at a 0.1 MOI and selected with 2 µg/ml puromycin (Gold Bio).

### 4.3 Lentivirus production

Lenti-X 293T cells were plated and transiently transfected with 1.1 µg of a GML plasmid library, 1.1 µg psPAX2, and 0.6 µg pMD2.G using FuGENE HD. Supernatant containing virus was harvested ∼72 h post-transfection, filtered through a 0.45 µM filter and viral titers were measured with Lenti-X GOSTIX (Takara).

### 4.4 GigaAssay Measuring the HER2 phosphorylation

All experimental methods were as previously described using the HER2 cDNA (accession number: NM_004448; NP_004439).[13,16] For the GigaAssay, 40 million cells were plated and 24 h after plating variant expression was stimulated by treatment of cells with 100 ng/mL doxycycline (Thermo Fisher Scientific) for 72 h. 50 ng/mL of doxycycline was spiked into media every 24 h to maintain the 100 ng/mL concentration through-out variant induction. After 72 h, cells were harvested by trypsinization, then immunostained for pHER2 (Y1248).

Cells were fixed in 4% formaldehyde, permeabilized in 90% methanol and blocked with Bovine Serum Albumin as previously described.[66] Samples were immunostained with primary monoclonal antibodies raised against human HER2 (rabbit mAb, Cell Signaling) and/or pHER2(Y1248) (mouse mAb, Thermo Fisher Scientific) and secondary antibodies conjugated to Alexa Fluor 647 (for HER2 immunostaining) or Alexa Fluor 488 (for pHER2 immunostaining) for control experiments. For the GigaAssay experiment, cells were immunostained with secondary antibodies conjugated to PE for HER2 immunostaining Fluor 647 for pHER2 immunostaining.

### 4.7 Flow cytometry

Individual cell lines and those transduced with GMLs were analyzed and sorted by flow cytometry. All analytical flow cytometry experiments were with a Sony SH800Z flow cytometer (SONY, Tokyo, Japan). Cells were first gated for high expression of HER2 (AF647) and then compared to their level of pHER2 (AF488). FACs data analysis for individual control experiments was performed using FlowJo (v. 10.8.1, Ashland, OR). Sorting of cell populations for the GigaAssay was performed by the Stanford flow Cytometry Core on a FACsAria II (BD, Franklin Lakes, NJ). Cells were gated for high expression of HER2 (PE) then sorted in 4 bins gated for increasing levels of pHER2 expression (AF647).

### 4.8 Bioinformatics

Each GigaAssay is assessed with long-read NGS sequencing of a UMI-barcoded plasmid variant library and short read sequencing of gDNA isolated from flow-sorted bins. The pipeline is designed to call variants for each UMI-barcoded cDNA and then calculate an activity score for each UMI-barcoded cDNA from UMI-barcode frequencies of short reads for each bin from cell sorting. The pipeline produces a UMI-barcode mutant map, which creates an index between UMI-barcodes identified in the long-read library and the mutation in the library. This index is used when flow-sorted short reads are produced and used for quantification of the UMI-barcodes to produce an activity score.

PacBio CCS long reads for the variant library and Illumina short reads for UMI-barcodes were filtered for low-quality reads using Trimmomatic.[67] Reads were trimmed with Cutadapt such that the variant region and UMI-barcode are within the read with a 15 BP buffer before and after the variant region.[68] Reads were trimmed with Cutadapt to remove excess BPs. Cutadapt detects, trims, and reverses reverse complement reads. UMI-barcodes of length 28-36 were extracted from reads and selected with the Cutadapt.[68] Sequences with UMI-barcodes were clustered with Starcode using a Levenshtein distance of 4 to avoid data loss due to sequencing errors.[69] Reads with the same UMI-barcodes will occur in bins sorted by flow cytometer. Reads that have the same UMI-barcode from different samples are grouped with a custom python script. The trimmed and filtered cDNAs are aligned to the reference sequence using BWA.[70,71] In order to index each UMI-barcode to a specific mutant, variants are called with a custom script. The resulting VCF file is associated with each UMI-barcode group. Insertion and deletion variants (indels) are identified with the program Nanocaller with parameters optimized for each library analyzed.[72]

After variant calling, a mutant-barcode map was created for interpretation of the short-read sorting gates. A custom python script was used to filter reads based on QC and a required read depth per UMI-barcode. When working with CCS reads, QC 30, and depth of 1 is recommended. Instead, the *ERBB2* library was processed with QC 0 minimum and depth of 1, with additional downstream filtration.

### 4.9 Statistics

Activity scores are calculated for each UMI-barcode by calculating the weighted mean of the read distributions of each UMI-barcode across the 4 flow sorted bins. UMI-barcodes are then grouped into distributions for each mutant, and WT. Right tailed t-tests compare each mutant’s distribution to WT’s distribution. Significant (P<0.05) mutants are classified as GOF mutants for the experiment.

### 4.10 Figure preparation

Most figures are prepared with matplotlib in python. Flow Cytometry profiles are created with FlowJo V5.00000. Cartoon diagrams are created with BioRender. Contacting residues in structures are calculated with default parameters of Ring 2.0 and use structure diagrams.[73] Structure surface and ribbon diagrams are made with PyMol.[74] Kaplan-Meier plots are created using the lifelines python library.[75] Many multi-panel figures are arranged with Lucid. Some heatmaps are generated with a Heligenics data portal; a description of this web application will be described in a separate paper. Other heatmaps are generated with Microsoft Excel.

## Data Availability

All data produced in the present work are contained in the manuscript.

## Acknowledgements.

We wish to thank for Stanford University Flow Cytometry Core for help with flow sorting of GigaAssay Samples and the Nevada Institute of Personalized Medicine Genome Acquisition and Analysis Core for NGS sequencing of GigaAssay samples. We acknowledge partial funding of this work by a grant award from the National Human Genome Research Institute, Grant number: R43HG012537. We thank Heligenics and Jackson Laboratories for partial funding this work.

## Supplementary Data

**Supplementary Figure 1.** GigaAssay variant scores for technical replicate 2

**Supplementary Figure 2.** GigaAssay variant classifications for technical replicate 2

**Supplementary Figure 3.** p values for HER2 variants for technical replicate 1

**Supplementary Figure 4.** p values for HER2 variants for technical replicate 2

**Supplementary Figure 5.** UMI-barcode counts for HER2 variants for technical replicate 1

**Supplementary Figure 6.** UMI-barcode counts for HER2 variants for technical replicate 2

**Supplementary Figure 7.** Read counts for HER2 variants for technical replicate 1

**Supplementary Figure 8.** Read counts for HER2 variants for technical replicate 2

**Supplementary Figure 9.** GOF variants that can be created from a 1 bp substitution

**Supplementary Figure 10.** Correlation between GigaAssay GOF mutant scores and survival

**Supplementary File 1.** GigaAssay variant and score replicate 1 dataset

**Supplementary File 2.** GigaAssay variant and score replicate 2 dataset

