## Supplementary Table for "New Gain-of-Function Mutations Prioritize Mechanisms of HER2 Activation"

# A

| GigaAssay | | Expected | |
|---|---|---|---|
| | | GOF | Non-GOF |
| Tested | GOF | 10 | 0 |
| | Non-GOF | 3 | 7 |
| | Inconclusive | 0 | 0 |

| MAVE | | Expected | |
|---|---|---|---|
| | | GOF | Non-GOF |
| Tested | GOF | 1 | 1 |
| | Non-GOF | 11 | 5 |
| | Inconclusive | 1 | 1 |

# B

| | F1 | Accuracy | Sensitivity | Specificity | PPV | NPV |
|---|---|---|---|---|---|---|
| GigaAssay | 0.87 | 0.85 | 0.77 | 1.00 | 1.00 | 0.70 |
| MAVE | 0.14 | 0.33 | 0.08 | 0.83 | 0.50 | 0.31 |

**Supplemental Table S1**. **A**. Comparison of mutant classifications for GigaAssay and MAVE experiments for HER2 variant effect. **B**. Comparison of method performance for GigaAssay and MAVE experiments for HER2 activity.
