## Supplementary Figures for "New Gain-of-Function Mutations Prioritize Mechanisms of HER2 Activation"

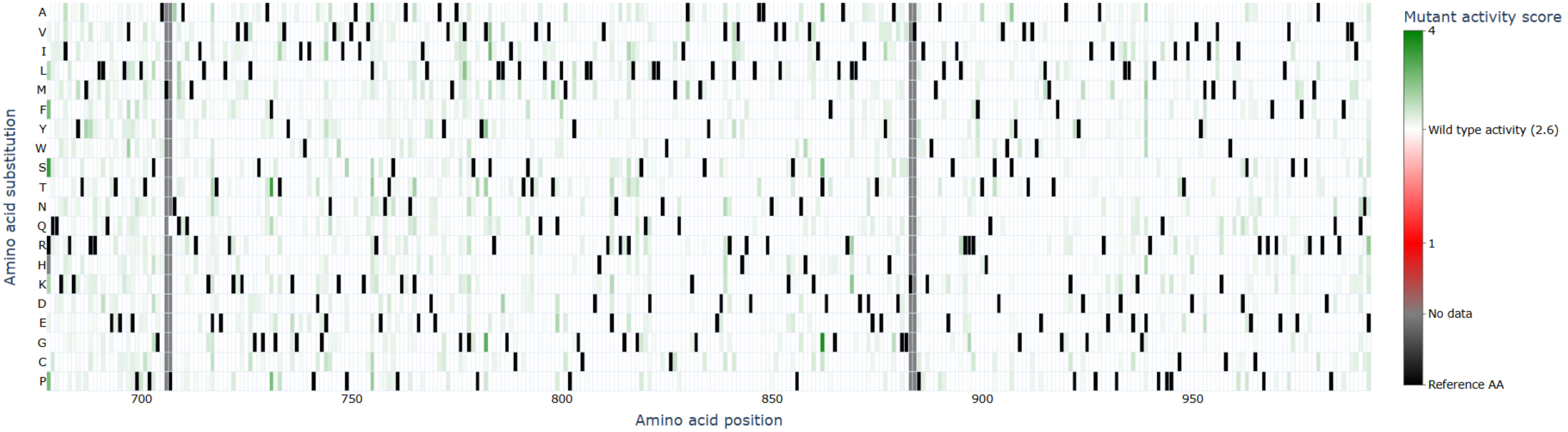


**Supplementary Figure S1: GigaAssay saturation mutagenesis mutant activity scores for replicate 2.** Each possible single amino acid substitution is represented by an individual box in the grid. The alternate amino acid is represented on the Y-axis, while the position is represented on the X-axis. The color of the cell indicates the functional activity of the mutation as indicated by the color key. Black squares are the reference sequence, and gray boxes have no data. The shade of green in the boxes indicate the level of functional activity of the HER2 mutant.

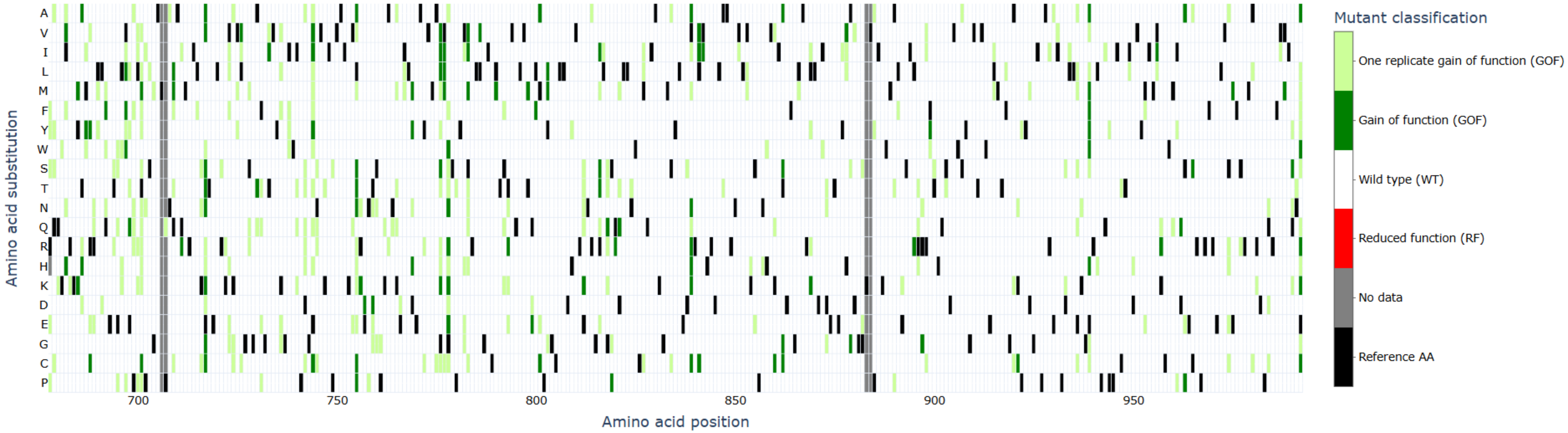


**Supplementary Figure S2: GigaAssay saturation mutagenesis mutant classifications for replicate 2.** Each possible single amino acid substitution is represented by an individual box in the grid. The alternate amino acid is represented on the Y-axis, while the position is represented on the X-axis. The color of the cell indicates the classification of the mutation as indicated by the color key. Black squares are the reference sequence, and gray boxes have no data.The color of the boxes indicates the classification of the mutants based upon statistical analysis. Data are from replicate 2.

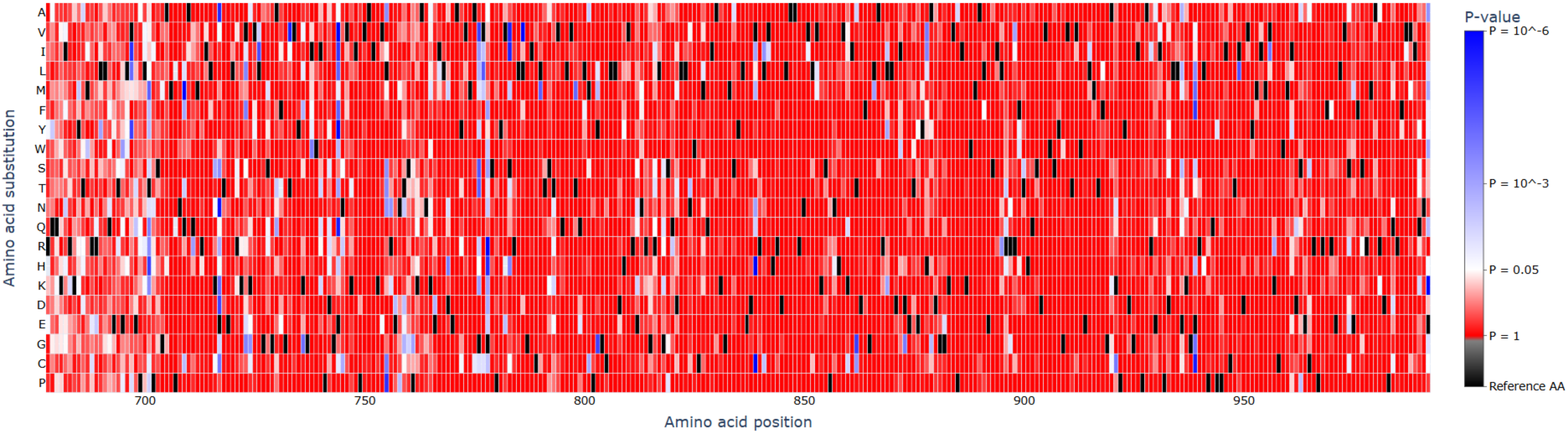


**Supplementary Figure S3: GigaAssay saturation mutagenesis p-values for replicate 1.** Each possible single amino acid substitution is represented by an individual box in the grid. The alternate amino acid is represented on the Y-axis, while the position is represented on the X-axis. The color of the cell indicates the p-value of the mutation activity score as indicated by the color key. Black squares are the reference sequence. The shade of blue in the boxes indicate the level statistical significance for the HER2 mutant being different than WT HER2. White boxes indicate mutants that are close to a threshold p-value of 0.05 and shades of red indicate not significantly different from WT HER2. Data are from replicate 1.

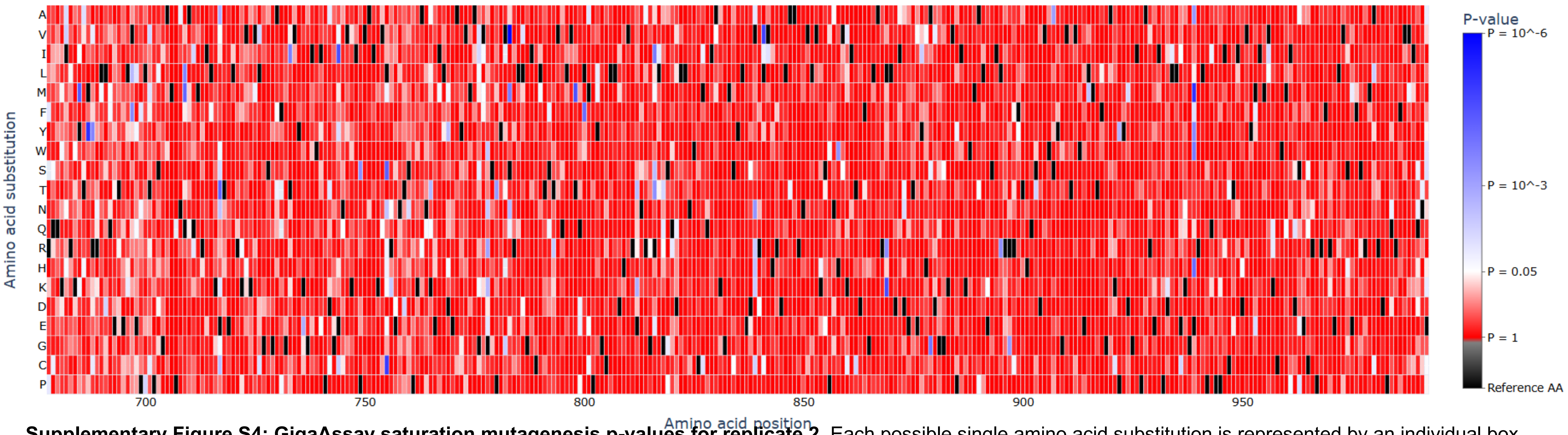


**Supplementary Figure S4: GigaAssay saturation mutagenesis p-values for replicate 2.** Each possible single amino acid substitution is represented by an individual box in the grid. The alternate amino acid is represented on the Y-axis, while the position is represented on the X-axis. The color of the cell indicates the p-value of the activity score for the mutation as indicated by the color key. Black squares are the reference sequence. The shade of blue in the boxes indicate the level statistical significance for the HER2 mutant being different than WT HER2. White boxes indicate mutants that are close to a threshold p-value of 0.05 and shades of red indicate not significantly different from WT HER2. Data are from replicate 2.

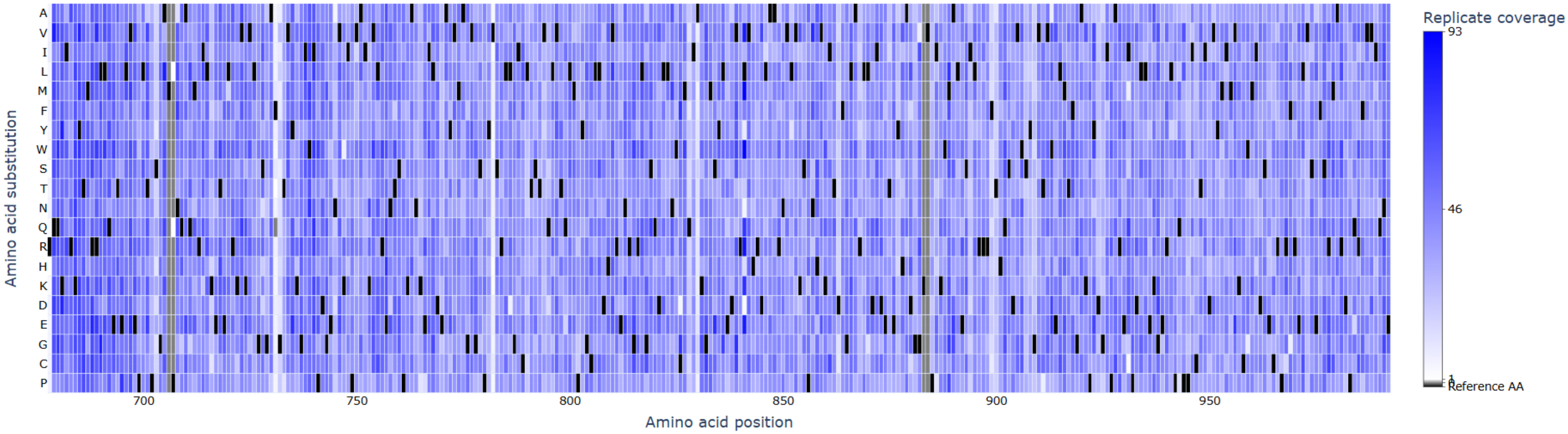


**Supplementary Figure S5: GigaAssay saturation mutagenesis number of barcodes for each mutant in replicate 1.** Each possible single amino acid substitution is represented by an individual box in the grid. The alternate amino acid is represented on the Y-axis, while the position is represented on the X-axis. The color of the cell indicates the the number of barcodes for the mutation as indicated by the color key. Black squares are the reference sequence, and gray boxes have no data. The shade of blue in the boxes indicate number of barcodes for the HER2 mutant.

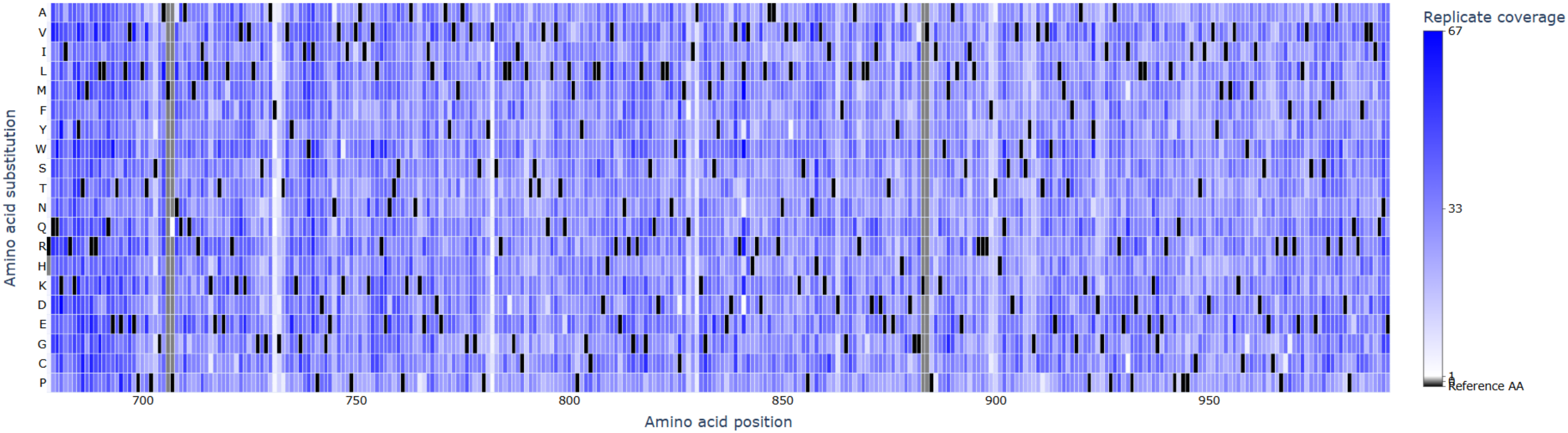


**Supplementary Figure S6: GigaAssay saturation mutagenesis number of barcodes for each mutant in replicate 2.** Each possible single amino acid substitution is represented by an individual box in the grid. The alternate amino acid is represented on the Y-axis, while the position is represented on the X-axis. The color of the cell indicates the the number of barcodes for the mutation as indicated by the color key. Black squares are the reference sequence, and gray boxes have no data. The shade of blue in the boxes indicate number of barcodes for the HER2 mutant.

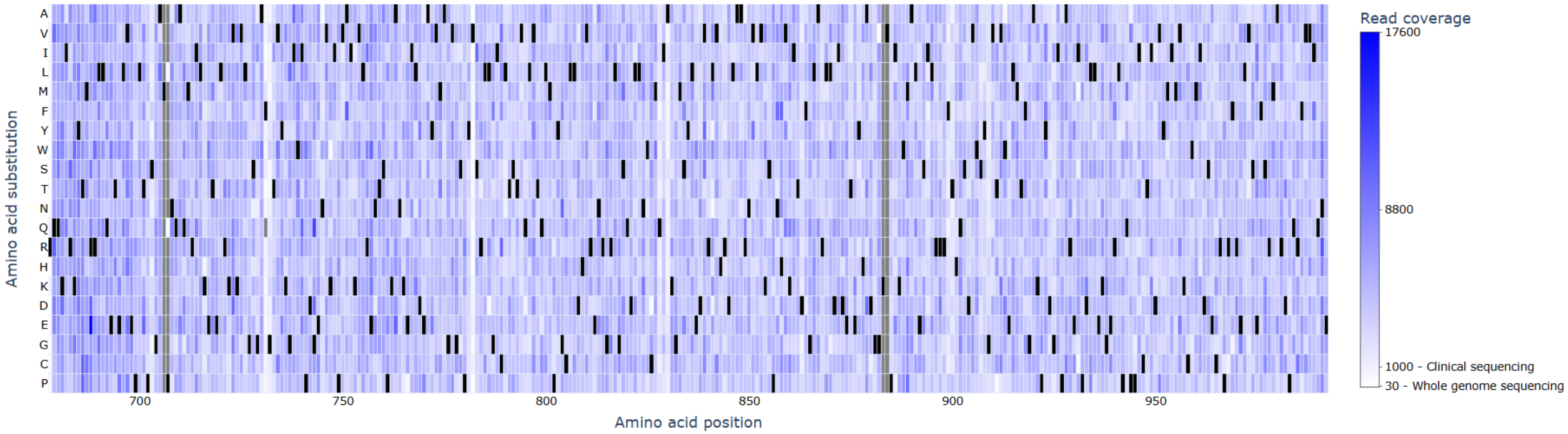


**Supplementary Figure S7: GigaAssay saturation mutagenesis number of reads for each mutant in replicate 1.** Each possible single amino acid substitution is represented by an individual box in the grid. The alternate amino acid is represented on the Y-axis, while the position is represented on the X-axis. The color of the cell indicates the the number of reads for the mutation as indicated by the color key. Black squares are the reference sequence, and gray boxes have no data. The shade of blue in the boxes indicate number of reads for the HER2 mutant.

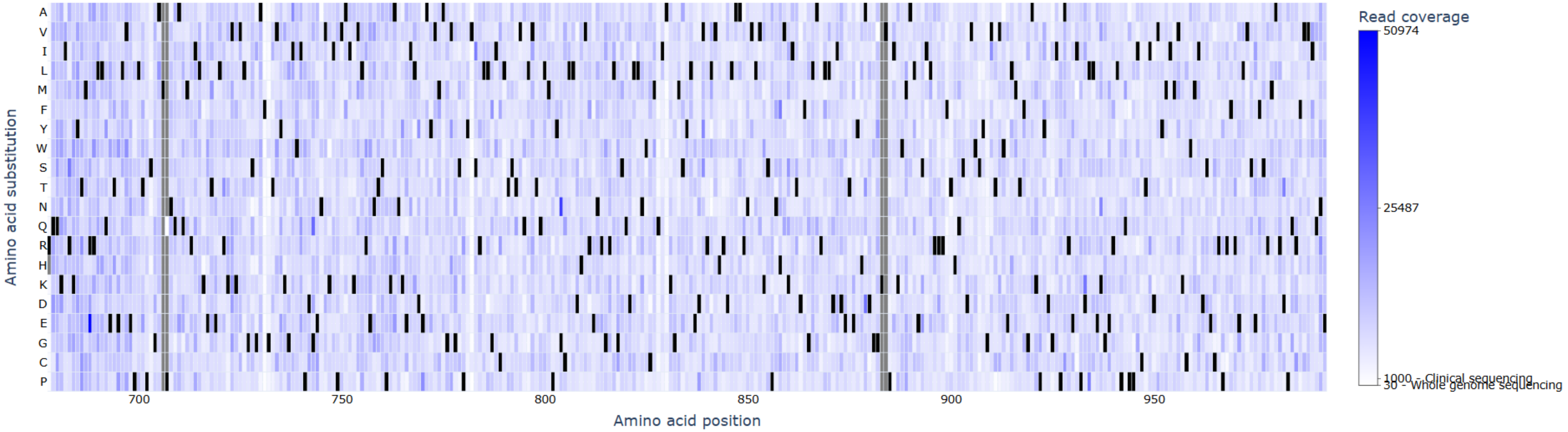


**Supplementary Figure S8: GigaAssay saturation mutagenesis number of reads for each mutant in replicate 2.** Each possible single amino acid substitution is represented by an individual box in the grid. The alternate amino acid is represented on the Y-axis, while the position is represented on the X-axis. The color of the cell indicates the the number of reads for the mutation as indicated by the color key. Black squares are the reference sequence, and gray boxes have no data. The shade of blue in the boxes indicate number of reads for the HER2 mutant.

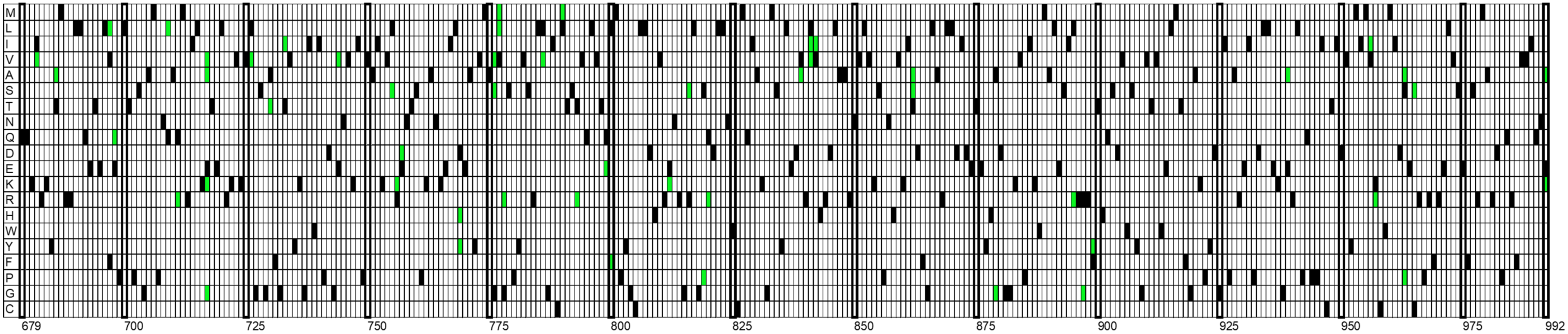


**Supplementary Figure S9: GigaAssay saturation mutagenesis mutant classification for substitutions that require only a 1-bp substitution.** Each possible single amino acid substitution is represented by an individual box in the grid. The alternate amino acid is represented on the Y-axis, while the position is represented on the X-axis. The green color of the cell indicates the the GOF mutations that would require only a 1-bp substation to for the mutation in the box.

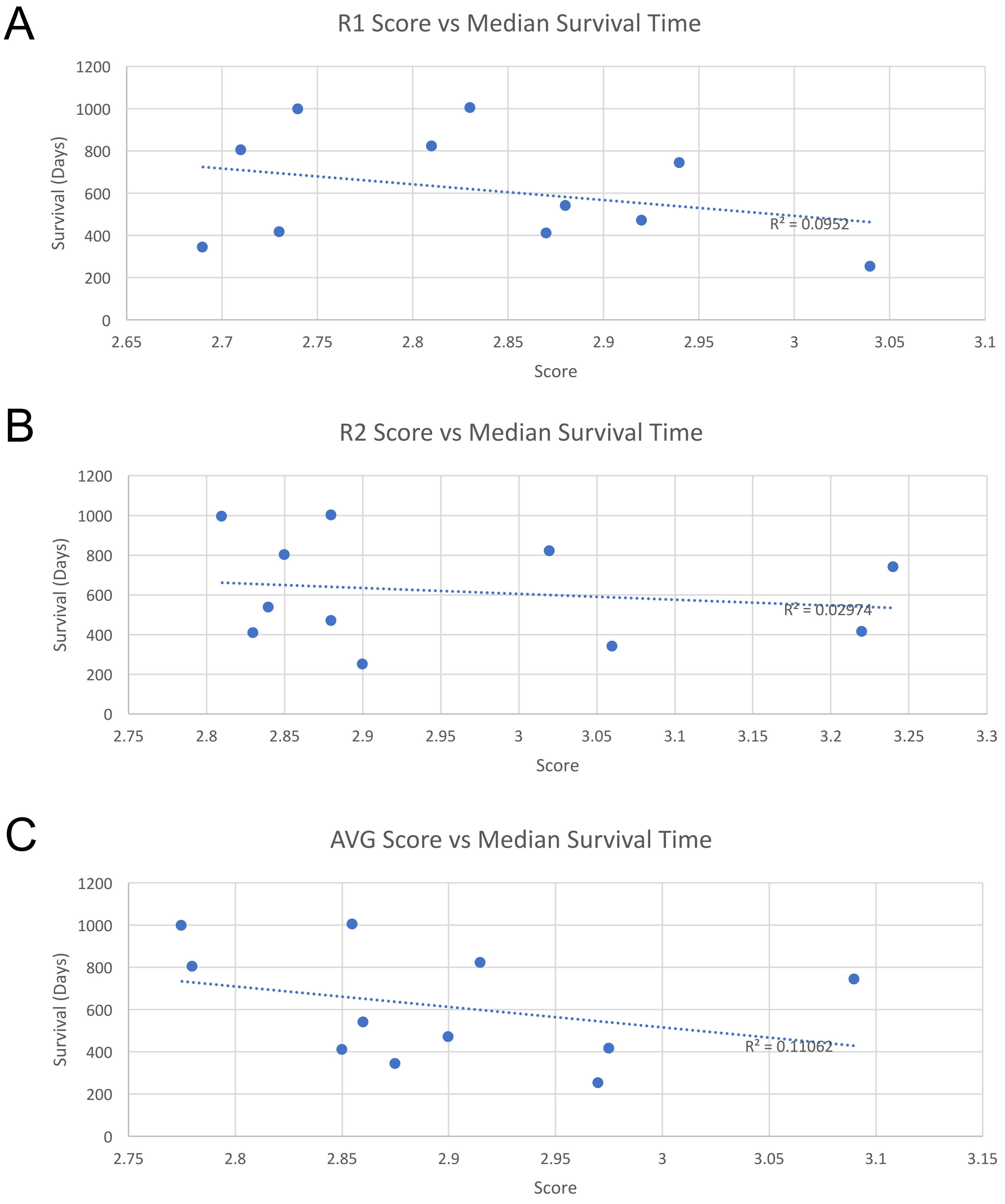


**Supplementary Figure S10: Relationship of HER2 hyperphosphorylation activity score from the GigaAssay to survival for cancer patients with a HER2 GOF. A, B, C.** The GigaAssay activity score was plotted against patient survival. A line was fit to the data and Pearson Correlation Coefficients are reported on each graph. ($R^2$ = 0.11). Data are for replicate 1 (**A**), replicate 2 (**B**), and averaged (**C**).
